# Microbial and chemical hazards in multi-use dryland waterpans: temporal variability, local heterogeneity and implications for drinking-water safety

**DOI:** 10.64898/2026.10.28.26366785

**Authors:** Sharon F. Akaranga, Edith J. Kurui, Kelvin O. Moenga, Claire Furlong, Jack van de Vossenberg, Frank O. Masese

## Abstract

Waterpans are widely used for domestic water supply, livestock, and other livelihood needs across drylands, yet spatial and temporal controls on their drinking-water quality remain poorly understood. We assessed 12 multi-use waterpans over nine sampling campaigns spanning dry and wet seasons, measured physicochemical variables, evaluated guideline-relevant chemical exposure, and tested for *Escherichia coli*. Water quality did not differ systematically among the three within-pan sampling positions, supporting analysis at the waterpan level. *E. coli* was detected in all 82 waterpan visits with microbiological data, indicating pervasive non-compliance with drinking-water standards. Fluoride exceeded 1.5 mg L⁻¹ in 36 of 58 dry-classified visits and 13 of 48 wet-classified visits, while the combined nitrate–nitrite guideline was exceeded in 24 of 106 visits, 23 of them during dry-classified campaigns. Mean child fluoride hazard quotients exceeded 1 at every waterpan in the dry season. Season was the strongest driver of physicochemical variation: eight of 14 variables (pH, total dissolved solids, conductivity, total phosphorus, nitrate, nitrite, fluoride and chemical oxygen demand) were higher in the dry season. Multivariate water quality differed among individual waterpans, whereas land use was not significant, although only 12 waterpans were available. *E. coli* varied mainly among sampling occasions and individual visits rather than persistently among waterpans. These findings indicate that drinking-water risk in dryland waterpans is multi-hazard and highly heterogeneous. Management should combine universal microbial protection and point-of-use treatment with targeted fluoride and nitrate–nitrite monitoring, intensified surveillance during low-water periods, and waterpan-specific interventions rather than relying on broad land-use classifications alone.

## 1. Introduction

Drylands cover about 40–45% of Earth’s terrestrial surface and support more than two billion people, yet low and highly variable rainfall, high evaporative demand, and recurrent drought constrain water availability in these landscapes (Huang et al., 2016; Prăvălie, 2016). Capturing episodic rainfall and runoff in small surface-storage systems is therefore an important component of water security, livestock production and climate adaptation across many dryland regions. These systems come in different forms and designs, including farm ponds and stock dams in Australia, tanks in South Asia, hafirs in northeastern Africa, small reservoirs in Mediterranean and Latin American drylands, and excavated or embanked waterpans in eastern Africa. Although these systems differ in origin and governance, many share a common functional template: they are relatively shallow, strongly coupled to their catchments, dependent on episodic runoff, subject to pronounced filling and drawdown cycles, and frequently used for several purposes simultaneously. Rainwater harvesting is consequently more than an engineering response to water scarcity; it creates small aquatic ecosystems whose hydrological reliability and water quality jointly determine their value to people and landscapes (Ritchie et al., 2021; Vohland & Barry, 2009).

Despite their abundance, research and management have disproportionately neglected small standing waters. Historically, research, monitoring programmes, and regulatory frameworks have focused on rivers, large lakes, and reservoirs, even though ponds and other small waters are among the most numerous freshwater habitats globally and contribute substantially to ecosystem functioning (Biggs et al., 2017; Downing, 2010). Their ecological significance extends well beyond their small individual area. In European agricultural landscapes, ponds can support greater regional richness of aquatic plants and macroinvertebrates, as well as more unique taxa and rarer species, than streams, ditches, and even larger waterbodies (Davies et al., 2008; Williams et al., 2004). Farm dams in Australia can act as reservoirs and stepping stones for aquatic biodiversity in otherwise modified landscapes (Brainwood & Burgin, 2009), while pond networks also support urban and peri-urban freshwater conservation, including habitat for threatened species (Oertli & Parris, 2019). Small ponds are also biogeochemically active because their shallow depths, extensive sediment–water interfaces and high perimeter-to-volume ratios promote intense nutrient processing and carbon exchange (Holgerson & Raymond, 2016). Thus, small waterbodies can simultaneously function as water infrastructure, biodiversity refugia and biogeochemical hotspots.

These functions are particularly sensitive to hydroperiod—the timing, duration, depth and spatial extent of inundation. Filling connects small waterbodies to their catchments through runoff and sediment transport, while drawdown progressively reduces volume, increases residence time and can concentrate dissolved constituents. Wetting and drying also alter oxygen conditions, nutrient transformations, microbial activity and habitat availability (Biggs et al., 2017; Céréghino et al., 2014). Where people, livestock, and wildlife access the same waterbody, these hydrological processes interact with direct faecal inputs, shoreline disturbance, sediment resuspension, and organic-matter loading. Consequently, the conditions that make small dryland waters valuable during periods of scarcity can also increase exposure to microbial and chemical hazards. Assessing such systems only as storage structures—or using a single indicator of water quality—can therefore obscure important trade-offs among water security, ecosystem condition and human health.

These trade-offs matter especially in African drylands, where small runoff-fed waterbodies support pastoral, agro-pastoral, and wildlife-dependent landscapes but remain poorly represented in long-term water-quality datasets. Studies of natural and constructed pans in southern Africa show that hydroperiod, salinity, vegetation, turbidity and disturbance structure biological communities and ecological condition (De Klerk et al., 2016; Masina et al., 2023; Nhiwatiwa et al., 2017). Rainwater harvesting is also widely promoted as a climate-adaptation and livelihood strategy across sub-Saharan drylands (Vohland & Barry, 2009). Yet multiple use can create strong human–livestock–wildlife interfaces, particularly where pans are unfenced, and users enter the water directly. Under these conditions, microbial contamination, nutrient enrichment and physicochemical deterioration may vary not only with rainfall and drawdown, but also among individual waterbodies according to morphology, catchment connectivity, sediment properties, intensity of use and management. Comparative evidence sufficient to distinguish these influences remains limited.

The East African Rift adds another chemical water-quality concern. Volcanic geology and alkaline hydrogeochemical conditions can generate elevated fluoride concentrations in groundwater and associated waters, with widespread exceedance of the 1.5 mg L⁻¹ drinking-water guideline reported from the Rift Valley (Gaciri & Davies, 1993; Olaka et al., 2016). Fluoride occurrence in surface waterpans cannot be assumed to have the same source without direct hydrogeochemical tracing, but the regional geochemical setting makes fluoride an important constituent to monitor alongside microbial contamination. Kenyan dryland studies have also reported variable microbial quality in waterpans and other unprotected surface-water sources (Kurui et al., 2017; Njarui et al., 2014). These hazards have different implications and controls: *Escherichia coli* indicates recent faecal contamination and failure of drinking-water microbiological safety, whereas fluoride and inorganic nitrogen represent chemical exposure pathways. Their co-occurrence therefore requires a multi-hazard, rather than single-parameter, assessment.

A central unresolved question is which spatial and temporal scales best explain water-quality variability in multi-use dryland waterbodies. Studies commonly invoke broad land-use classifications because agriculture, livestock, and wildlife influence contaminant delivery, but these categories may mask substantial heterogeneity among individual waterbodies. Conversely, repeated observations from the same pan may vary strongly among sampling campaigns as water levels, antecedent rainfall and use intensity change. Evidence from a regulated Afrotropical reservoir cascade showed that temporal factors explained considerably more water-quality variation than spatial position or land use (Ibrahim et al., 2026), although deep reservoirs and shallow waterpans differ fundamentally in hydrology and disturbance. For small, runoff-fed pans, the relative contributions of campaign-level hydrological conditions, individual-waterpan identity, surrounding land use and morphology remain poorly resolved. Distinguishing these scales has practical consequences: temporal dominance would favour strategically timed monitoring; consistent among-pan differences would require site-specific management; and strong land-use effects would support interventions organised at the catchment or landscape scale.

We addressed these questions using repeated observations from 12 multi-use waterpans in semi-arid rangelands in Kenya, sampled on nine campaigns between November 2024 and February 2026. We sampled each waterpan (pan) at three functionally distinct positions associated with human water collection, livestock access, and comparatively undisturbed water, allowing us to first test whether within-pan use zones required separate treatment before analysing waterpan-level patterns. The study then pursued four objectives: (i) to characterise microbiological and physicochemical drinking-water quality and the frequency of guideline-relevant exceedances; (ii) to quantify campaign-level dry–wet variation while accounting explicitly for repeated measurements of the same waterpans; (iii) to determine the extent to which water quality varies among individual waterpans and broad land-use classes, and to evaluate whether measured morphology helps explain among-pan differences; and (iv) to characterise microbial non-compliance and screening-level chemical exposure, particularly from fluoride and the combined occurrence of nitrate and nitrite.

We tested three principal expectations. First, because the pans are open, intensively used and shared among humans, livestock and wildlife, we expected faecal contamination to be widespread across land-use classes rather than confined to a small subset of pans. Second, we expected dry-season campaigns to show higher concentrations of conservative dissolved constituents and inorganic nitrogen because reduced dilution and declining water volume favour concentration, while recognising heterogeneity in individual water quality responses. Third, we expected substantial, persistent variability among individual waterpans that broad land-use categories would not fully capture, because local morphology, hydroperiod, sediment characteristics, access patterns, and management differ among pans. Together, these tests shift the emphasis from describing water quality at a single locality to evaluating how multi-use dryland waterbodies function as a class of small freshwater systems and at which spatial and temporal scales their drinking-water risks should be monitored and managed.

## 2. Materials and Methods

### 2.1. Study area

The study was conducted in 12 waterpans distributed along the Mulot-Narok and Narok-Sekenani transects in Narok County, southeastern Kenya (Figure 1). The waterpans occur within the semi-arid rangelands at elevations ranging from 1753–2039m above sea level. Land use is dominated by grasslands and shrublands, but large areas are increasingly under cultivation. The rainfall is bimodal and influenced by the inter-tropical convergence zone. The long rains are experienced from March to June while the short rains occur from October to December (Narok County Government, 2023); higher-elevation areas receive substantial rainfall outside these periods (MEMR, 2009; Olang, 2025; Tuna, 2025). Within the study area, rainfall varies with elevation, ranging from 1000 mm in higher-elevation areas and 400 mm in lowlands (GoK, 2013). The region encompasses ecologically important savanna, rangeland, and wetland habitats that support diverse wildlife communities, including large herbivore populations (Ottichilo et al., 2000). Pastoral and agro-pastoral livelihoods are widespread, and constructed waterpans provide important water for domestic use, livestock, wildlife and small-scale agriculture (Narok CIDP, 2018). Most pans are therefore embedded within a shared human–livestock–wildlife water-use interface in which direct access, catchment runoff and seasonal drawdown can jointly influence water quality.

**Fig. 1.**
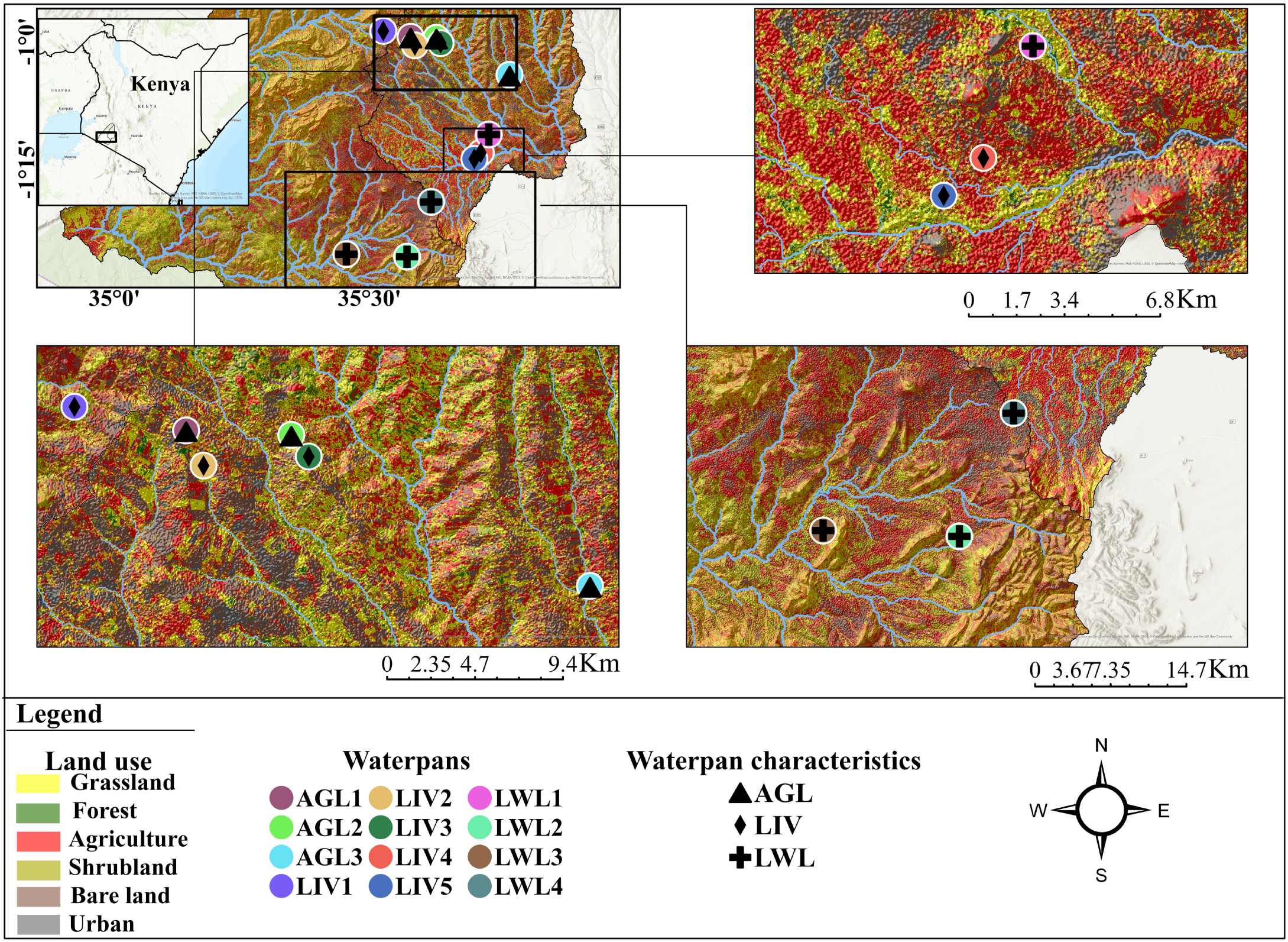
Location of the 12 waterpans sampled along the Mulot–Narok and Narok–Sekenani transects in Narok County, Kenya. Colours identify individual waterpans and symbols the land-use category (triangle, agriculture–livestock, AGL; diamond, livestock, LIV; cross, livestock–wildlife, LWL). The background shows land cover

The geological and hydrogeological setting is particularly relevant to dissolved-ion and fluoride dynamics. Narok County lies within the southern Kenya Rift and contains a heterogeneous mixture of Precambrian Mozambique Belt basement rocks and younger Rift-related volcanic formations. Quartzites and muscovite quartzites are prominent around the Loita Hills, whereas extensive lower-lying areas are underlain by phonolites, trachytes and basalts interbedded with volcanic ash and tuffs (Tuna, 2025; Wright, 1967). These lithologies create contrasting groundwater-flow pathways: fractured quartzites and basalts can promote infiltration, recharge and groundwater movement, whereas less-permeable phonolitic and trachytic units may concentrate flow along fractures, faults and lithological contacts where springs emerge (Saggerson, 1966; Tuna, 2025). Hydrochemical and isotopic work in the wider Narok waterpan system further indicates that pans range from predominantly rainfall-and runoff-fed systems to systems receiving groundwater or spring contributions. Tuna (2025) identified Na–HCO₃–F–Mn signatures associated with deeper groundwater and showed that spatial variation in volcanic materials, quartzitic outcrops and groundwater connectivity contribute to differences in waterpan chemistry. This setting provides plausible geogenic pathways for fluoride because elevated fluoride is well documented in alkaline volcanic waters of the East African Rift, where water–rock interaction and prolonged residence time can promote fluoride mobilisation (Gaciri & Davies, 1993; Olaka et al., 2016). Evaporation from shallow, seasonally contracting pans may subsequently concentrate dissolved constituents.

### 2.2. Study and sampling design

Purposive sampling was used to select 12 waterpans used by local communities for drinking water supply. The selected waterpans were classified into three land use categories based on the dominant land use within their surrounding catchments and the number of water users associated with each waterpan: agriculture and livestock (AGL), livestock (LIV), and livestock and wildlife (LWL). The AGL category comprised waterpans located within agro-pastoral landscapes where livestock production co-occurred with crop cultivation. The LIV waterpans were mainly located in pastoral areas characterised by livestock grazing with minimal cultivation, whereas the LWL category consisted of waterpans shared by livestock and free-ranging wildlife within pastoral rangelands adjacent to wildlife conservancies and dispersal areas. Accordingly, we classified three waterpans as AGL, five as LIV, and four as LWL (Table 1; Figure 1). The 12 waterpans comprised six sites along the Mulot-Narok transect and six along the Narok-Sekenani transect. Sampling was conducted on nine occasions between November 2024 and February 2026 to capture temporal variation in water quality across the wet and dry seasons. Four occasions fell within the wet season (January 2025, May 2025, June 2025, and February 2026), and five fell within the dry season (November 2024, October 2025, November 2025, December 2025 and January 2026). January was sampled under both conditions in different years, illustrating this variability. The dry season classification of December 2025 and January 2026 is supported by LWL1 drying out completely on both occasions.

**Table 1.** Location of sampled waterpans, their characteristics and classification based on land use.

| Waterpan code | Local name | Latitude (°) | Longitude (°) | Mean depth (Wet season) (m) (n=10) | Mean depth (Dry season) (m) (n=10) | Mean area (Wet season) (m<sup>2</sup>) (n=12) | Mean area (Dry season) (m<sup>2</sup>) (n=12) | Shape | Land use |
| --- | --- | --- | --- | --- | --- | --- | --- | --- | --- |
| LIV1 | Longisa | -0.97860 | 35.52700 | 1.80 | 1.30 | 6520.35 | 5939.75 | Irregular | Livestock (LIV) |
| AGL1 | Iltirben | -0.98995 | 35.58059 | 2.20 | 1.80 | 10379.1 | 10369.22 | Irregular | Agriculture and livestock (AGL) |
| LIV2 | Olonini | -1.00686 | 35.58898 | 1.86 | 1.60 | 578.57 | 506.60 | Square | Livestock (LIV) |
| AGL2 | Olarien | -0.99212 | 35.63109 | 1.60 | 0.54 | 1956.07 | 1676.78 | Rectangular | Agriculture and livestock (AGL) |
| LIV3 | Melelo | -1.00251 | 35.63948 | 1.20 | 1.19 | 428.83 | 318.75 | Square | Livestock (LIV) |
| AGL3 | Katakala | -1.06461 | 35.77451 | 1.90 | 1.30 | 623.98 | 455.80 | Rectangular | Agriculture and livestock (AGL) |
| LWL1 | Ewasonyiro | -1.18169 | 35.73399 | 1.00 | 0.34 | 3643.00 | 1506.02 | Square | Livestock and wildlife (LWL) |
| LIV4 | Majimoto | -1.21739 | 35.71819 | 2.90 | 2.00 | 5106.06 | 3601.78 | Square | Livestock (LIV) |
| LIV5 | Esupetai | -1.22942 | 35.70552 | 3.50 | 1.60 | 2461.65 | 1141.35 | Square | Livestock (LIV) |
| LWL2 | Ngoswani | -1.42119 | 35.57353 | – | – | 6802.22 | 6799.10 | Rectangular | Livestock and wildlife (LWL) |
| LWL3 | Kishermoruak | -1.41616 | 35.45466 | – | – | 4796.33 | 4335.05 | Rectangular | Livestock and wildlife (LWL) |
| LWL4 | Empura | -1.31374 | 35.62090 | 3.50 | 3.00 | 11007.32 | 9489.10 | Rectangular | Livestock and wildlife (LWL) |

NB: Depth could not be measured at LWL2 and LWL3 owing to the presence of hippopotamuses.

### 2.3. Sample collection

Within each waterpan, three fixed sampling points (A, B and C) were established to capture variation linked to different usage patterns: Point A at the human water-collection point, Point B at the livestock watering point, and Point C at an undisturbed area with minimal human or animal activity. The same points were revisited on each sampling occasion in the wet and dry seasons, and all samples were collected at 30 cm depth below the water surface. Where water levels had receded in the dry season, samples were collected at a depth of less than 30 cm. Because the three points reflected different patterns of use, we first tested whether water quality differed among them before any further analysis (Table S1).

Samples for total phosphorus (TP; mg L⁻¹), fluoride (F⁻), and chemical oxygen demand (COD) were collected in 500 mL high-density polyethylene (HDPE) bottles, with one sample at each sampling point (A, B and C). Samples were stored in a cooler box at 4 °C and transported to the laboratory for analysis. The samples for dissolved nutrient analysis were filtered through GF/F filters (Whatman, 0.70 µm pore size, 47 mm diameter), and the filtrate was stored in 30 mL and 60 mL HDPE bottles at 4 °C and transported to the laboratory for analysis of soluble nutrient fractions: soluble reactive phosphorus (SRP; µg L⁻¹), soluble ammonium (NH₄⁺; µg L⁻¹), soluble nitrate (NO₃⁻; mg L⁻¹), and soluble nitrite (NO₂⁻; mg L⁻¹).

Water samples for *Escherichia coli* were collected in 500 mL HDPE bottles that had been sterilised with sodium hypochlorite solution, rinsed with distilled water, air-dried, and pre-rinsed on-site with sample water prior to collection. Bottles were capped, stored in ice-filled cooler boxes at approximately 4 °C, and analysed either in the field or within 1–24 h of collection.

### 2.4. Field and laboratory analyses

#### 2.4.1. Physico-chemical variables

Water temperature (°C), pH, electrical conductivity (EC; µS cm⁻¹), dissolved oxygen concentration (DO; mg L⁻¹), oxygen saturation (OS; %), and total dissolved solids (TDS; mg L⁻¹) were measured *in situ* using a WTW Multi 3630 IDS portable multi-parameter probe (WTW GmbH, Weilheim, Germany) at three sampling points in each of the 12 waterpans. Water transparency and depth were determined using a Secchi disk and a hand-held echo sounder (Lubembe et al., 2024).

Water samples were analysed for fluoride, chemical oxygen demand (COD), and nutrient concentrations using standard methods (APHA, 2017). Fluoride was determined using an ion-selective electrode method (APHA 4500-F⁻ C). Chemical oxygen demand was determined using the closed reflux, colorimetric method (APHA 5220 D). Soluble reactive phosphorus (SRP) was determined using the ascorbic acid method (APHA 4500-P E), while total phosphorus (TP) was determined by persulfate digestion (APHA 4500-P B) followed by colorimetric determination using the ascorbic acid method (APHA 4500-P E). Soluble nitrite (NO₂⁻) was determined using the colorimetric diazotization method (APHA 4500-NO₂⁻ B), and soluble nitrate (NO₃⁻) was determined using the cadmium reduction method (APHA 4500-NO₃⁻ E). Soluble ammonium (NH₄⁺) was determined using the phenate method (APHA 4500-NH₃ F). Quality control followed APHA (2017): the multi-parameter probe was calibrated according to the manufacturer’s instructions, samples were kept at 4 °C during storage and transport, and fluoride, COD and nutrient concentrations were determined against calibration standards in line with the quality-control requirements of each method.

#### 2.4.2. Microbiological variable

Microbiological analysis was performed using the Faircap Portable Lab *E. coli* testing kit (Faircap CIC, London, UK). For each sample, two 100 mL duplicate water samples were each passed through a 0.45 µm pore size membrane filter using the kit’s syringe-driven concentrating disk, and each filter was transferred onto a Compact Dry EC (Shimadzu Diagnostics Corporation, Japan) chromogenic plate that was pre-wetted with 1 mL of sterile water. Plates were incubated in a Faircap portable incubator at 38–42 °C for 14–16 h, following the manufacturer’s protocol (Faircap CIC, 2022). Characteristic blue colonies were enumerated as *E. coli* and expressed as colony forming units (CFU) per 100 mL, corresponding directly to the 100 mL volume filtered. Slight clogging occasionally occurred, but we filtered the full 100 mL for all samples. The duplicate 100 mL filtrations of each sample provided a check on the precision of the *E. coli* counts.

### 2.5. Descriptive statistics and guideline comparison

Descriptive statistics (mean, standard deviation, median and range) were calculated for each of the 15 water quality parameters, grouped by season, land use and waterpan. Nine of the 15 parameters have an applicable World Health Organization (WHO, 2022) or Kenya Bureau of Standards (KEBS, 2014) drinking water guideline value and were compared against it, whereas the remaining six have no numerical drinking water guideline under either body and were retained as descriptive ecological and organic enrichment indicators (Tables S2 and S3). The proportion of samples exceeding each guideline value was calculated separately for the dry and wet seasons, with 95% confidence intervals where appropriate. For *E. coli*, compliance required no detectable count in 100 mL.

### 2.6. Data analyses

Each of the 14 physicochemical parameters was examined for skewness and heterogeneity of variance before analyses. Distributions were assessed with the Shapiro–Wilk test on raw and log₁₀-transformed values at α = 0.05. Ten parameters (TDS, EC, SRP, TP, soluble nitrate, soluble ammonium, soluble nitrite, fluoride, COD and Secchi depth) were strongly right-skewed on the raw scale and were log₁₀-transformed prior to analysis. Temperature, pH, dissolved oxygen and oxygen saturation were analysed untransformed. pH was not transformed since it is already a logarithmic measure of hydrogen-ion activity. Coefficients for the log₁₀-transformed parameters are reported on the log₁₀ scale. All analyses were performed in R version 4.5.2 (R Core Team, 2025), with data handling in dplyr, and figures produced with ggplot2 using ggrepel and patchwork extensions.

Analyses used two nested levels of averaging to avoid pseudo-replication. The first collapsed the three within-waterpan sampling positions (A, B and C) into a single value per waterpan per sampling occasion (n = 106 waterpan-occasion observations for the 14 physicochemical parameters; n = 82 for *Escherichia coli*, which was measured on fewer campaigns); this waterpan-occasion resolution is retained in sections 2.6.2 and 2.6.4, where waterpan is instead fitted as a random intercept to account for the repeated sampling of the same waterpans over time without discarding that temporal replication. The second level collapsed the waterpan-occasion values further, to a single seasonal mean per waterpan (n = 12 per season), and was used only where an analysis required one row per waterpan per season: the principal component analysis in section 2.6.3, and the fluoride exposure summaries in section 2.6.5.

#### 2.6.1. Within-waterpan sampling design and test of homogeneity

Each of the 12 waterpans was sampled at three fixed points (A, B and C) on every visit. Each campaign was classified as dry or wet from the rainfall reported by households using the waterpans in the 1–2 weeks before sampling, not from the long-term seasonal calendar described in Section 2.1, so the classification does not always coincide with the climatological seasons (Table S4). Of 324 planned observations (12 waterpans × 3 positions × 9 occasions), 318 were obtained because one waterpan (LWL1) was dry on two occasions. Sampling dates and visit-specific observations are provided in Table S4. *E. coli* was measured on seven of the nine occasions, as it was not included during the first two sampling campaigns (November 2024 and January 2025). This gave 246 of 252 planned observations (12 × 3 × 7) again after the two visits when LWL1 was dry. Because three fixed sampling positions were sampled within each waterpan during each visit, and the same waterpans were repeatedly sampled over time, observations were not independent. Before analysing seasonal, land use, and between-waterpan variation, we tested whether water quality differed among the three sampling positions (A, B, and C). For each physicochemical parameter, a linear mixed-effects model (LMM) was fitted with sampling position (A, B and C) as the fixed effect and random intercepts accounting for waterpan, sampling occasion and individual waterpan visit, so that the effect of position was tested against within-visit variation:

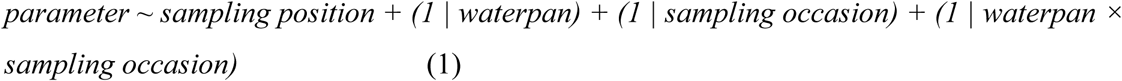

*E. coli* counts were overdispersed relative to the Poisson distribution (Pearson dispersion ratio = 111.65) and were therefore analysed using a negative-binomial generalised linear mixed-effects model with the same random-effects structure. The NB1 parameterisation (variance increasing linearly with the mean) was selected over NB2 (variance increasing quadratically with the mean) on the basis of Akaike’s information criterion (AIC) at the individual sample level (ΔAIC = 12.73). Within-pan position effects were assessed using F-tests with Kenward–Roger degrees of freedom for the linear mixed models and a Type II Wald χ² test for the *E. coli* model. Benjamini–Hochberg correction (Benjamini & Hochberg, 1995) was applied across the 15 sampling-position tests. Because a non-significant difference does not establish equivalence, pairwise equivalence among the three sampling positions was additionally assessed using two one-sided tests (TOST; Lakens, 2017), with an equivalence margin of 0.5 SD of the modelled response. Effect size was expressed as the largest pairwise difference in SD units (Table S1).

#### 2.6.2. Spatial and temporal variation in physicochemical water quality

Because water quality did not differ among the three within-waterpan positions (A, B and C), their readings were averaged to obtain a single value per waterpan per sampling occasion prior to modelling. Spatial and temporal variation in each parameter was assessed using linear mixed models (LMMs) fitted by restricted maximum likelihood in lme4 (Bates et al., 2015), with season and land use as fixed factors, waterpan area as a fixed covariate, and waterpan fitted as a random intercept to account for repeated sampling of the same waterpans over time:

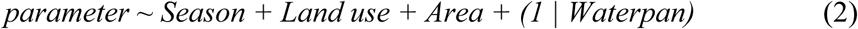

Depth was excluded from this model due to not being measured in LWL2 and LWL3 (Table 1) allowing the full sample of n = 106 waterpan-occasion observations from all 12 waterpans across nine campaigns (five dry season and four wet season). Depth was tested separately in a model of identical structure, fitted to the 10 waterpans where it was recorded (parameter ∼ Season + Land use + Area + Depth + (1 | Waterpan) (a variant of Equation 2 with Depth added), and reported as a sensitivity analysis (Table S5). Although the model was fitted to n = 106 waterpan-occasion observations, land use and waterpan area are properties of the waterpan rather than of the sampling occasion, so the effective replication available to test these two terms is the 12 waterpans, not the 106 observations; the season term, by contrast, is replicated within every waterpan across nine campaigns and is not subject to this limitation.

The wet season and the agriculture-livestock category were the reference levels, so season coefficients express the dry-season contrast directly. Fixed effect significance was assessed using t-tests with Satterthwaite-approximated denominator degrees of freedom in lmerTest (Kuznetsova et al., 2017). To test whether the seasonal direction was consistent across waterpans, dry and wet season means were also compared within each waterpan. Model performance was summarised using AIC and Nakagawa’s marginal and conditional R² (Nakagawa & Schielzeth, 2013), calculated with the performance package (Lüdecke et al., 2021).

#### 2.6.3. Multivariate analysis of water quality composition

Patterns in overall physicochemical water quality were visualised using principal component analysis (PCA) of 15 centred and standardised parameters, comprising 14 physicochemical parameters and *E. coli*. We imputed a small number of missing values using regularised iterative PCA, selecting the number of components by cross-validation (missMDA; Josse & Husson, 2016). PCA was used for visualisation only. Each waterpan was represented by its seasonal mean, and we computed separate PCAs for the dry and wet seasons (n = 12 waterpans per season). Seasonal *E. coli* means were based only on visits on which it was measured. We tested differences in overall composition using permutational multivariate analysis of variance (PERMANOVA; Anderson, 2001) based on Euclidean distances calculated from the 14 centred and standardised physicochemical parameters. *E. coli* was excluded because it was measured in only seven of the nine campaigns. We assessed the effects of waterpan, season, land use, and the season × land use interaction while accounting for repeated sampling of the same waterpans (Table S6). For waterpan, we restricted permutations within campaign. For season, we restricted permutations within waterpan so we could compare wet and dry season conditions within the same waterpan, and we also tested season across all 126 possible wet/dry assignments of the nine campaigns. We assessed land use and the season × land use interaction at the waterpan level because land use was a fixed characteristic of each waterpan, yielding 12 independent waterpans. Differences in multivariate dispersion between groups were assessed using betadisper and permutest (Table S7). PERMANOVA was performed using adonis2 in the vegan package (Oksanen et al., 2022), with 9,999 permutations and α = 0.05.

#### 2.6.4. Drivers of *Escherichia coli* concentrations

*Escherichia coli* counts were modelled using a negative-binomial generalised linear mixed model (NB1, log link), fitted by maximum likelihood in *glmmTMB* (Brooks et al., 2017).

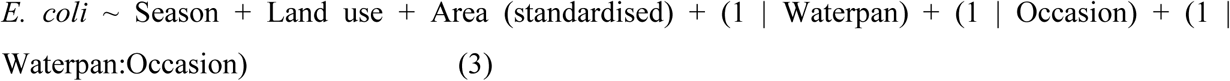

Waterpan area was the seasonal mean area of each waterpan, standardised as a z-score before fitting. A seasonal mean was used because area changed markedly between seasons. LWL1, for example, fell from 3,643 m² in the wet season to 1,506 m² in the dry. The wet season and the agriculture–livestock category were the reference levels. The model was fitted to the counts from the three within-pan positions, each the mean of duplicate plates rounded to the nearest whole number (n = 246, nested within 82 waterpan–occasion combinations). Effects are reported as incidence rate ratios (IRR) with 95% Wald confidence intervals, and the land-use term was tested with a joint Type II Wald test (car package; Fox & Weisberg, 2019). The full model was compared with a random-intercepts-only null model by AIC and a likelihood-ratio test (Table S8). As with the physicochemical models, land use and area are waterpan-level covariates fitted alongside the waterpan random intercept, so despite the 246 individual samples and 82 waterpan-occasion combinations used to fit the model, the effective replication for these two terms remains 12 waterpans.

#### 2.6.5. Fluoride exposure and non-carcinogenic hazard

Health risk was characterised at screening level for fluoride, assuming exposure through drinking water from waterpans. Chronic daily intake (CDI) of fluoride through drinking water was estimated using the standard United States Environmental Protection Agency (USEPA) exposure-assessment framework (USEPA, 1989a, 2011):

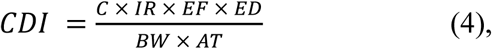

Where:

C is the fluoride concentration (mg L⁻¹),

IR the ingestion rate (L day⁻¹),

EF the exposure frequency (365 days year⁻¹, the USEPA screening default assuming daily consumption)

ED the exposure duration, BW the body weight (kg)

AT the averaging time (ED × 365 days)

EF = 365

AT = ED × 365,

Equation 4 simplifies to CDI = C × IR / BW.

The hazard quotient (HQ) was calculated as:

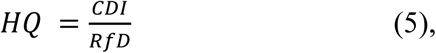

where RfD is the USEPA oral reference dose for dental fluorosis (USEPA, 1989b). An HQ above 1 indicates intake above the reference dose and therefore a potential non-carcinogenic risk. Table 2 gives exposure parameters for adults and children. We calculated CDI and HQ for each of the 318 individual samples and summarised them as arithmetic means per waterpan and season, along with the maximum sample-level HQ and the percentage of samples with HQ > 1 (Table S9). Under these assumptions, HQ=1 corresponds to a fluoride concentration of 0.9 mg L⁻¹ for children and 2.1 mg L⁻¹ for adults, and at the WHO and KEBS guideline value of 1.5 mg L⁻¹, the HQ is 1.67 for children and 0.71 for adults. Values pooled across seasons are given in Table S10.

**Table 2.**
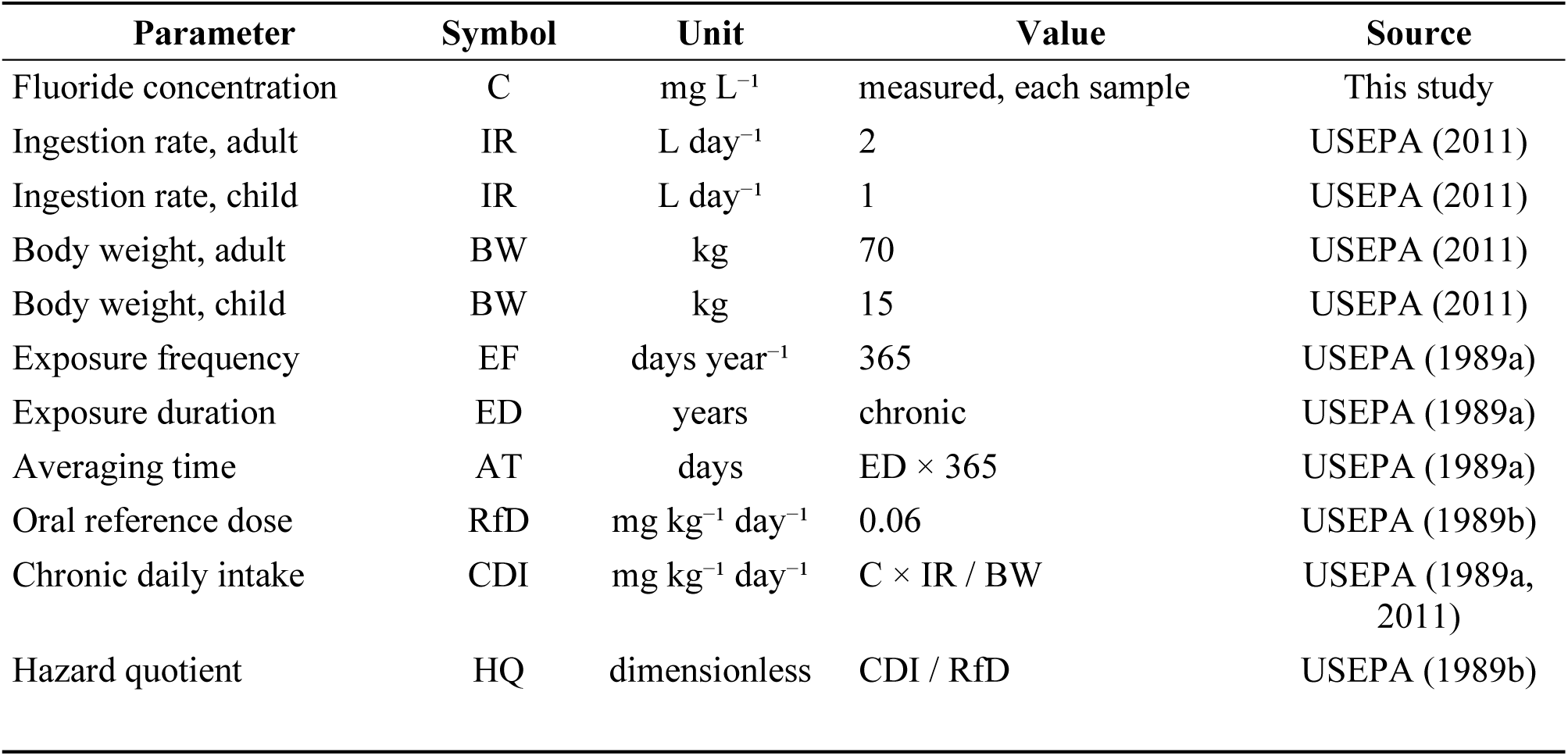
Exposure parameters used in the chronic daily intake and fluoride hazard quotient calculations.

| Parameter | Symbol | Unit | Value | Source |
| --- | --- | --- | --- | --- |
| Fluoride concentration | C | mg L<sup>-1</sup> | measured, each sample | This study |
| Ingestion rate, adult | IR | L day<sup>-1</sup> | 2 | USEPA (2011) |
| Ingestion rate, child | IR | L day<sup>-1</sup> | 1 | USEPA (2011) |
| Body weight, adult | BW | kg | 70 | USEPA (2011) |
| Body weight, child | BW | kg | 15 | USEPA (2011) |
| Exposure frequency | EF | days year<sup>-1</sup> | 365 | USEPA (1989a) |
| Exposure duration | ED | years | chronic | USEPA (1989a) |
| Averaging time | AT | days | ED × 365 | USEPA (1989a) |
| Oral reference dose | RfD | mg kg<sup>-1</sup> day<sup>-1</sup> | 0.06 | USEPA (1989b) |
| Chronic daily intake | CDI | mg kg<sup>-1</sup> day<sup>-1</sup> | C × IR / BW | USEPA (1989a, 2011) |
| Hazard quotient | HQ | dimensionless | CDI / RfD | USEPA (1989b) |

#### 2.6.6. Faecal exposure characterisation

We assigned each individual sample a WHO (1997) faecal risk category based on its *E. coli* concentration: in conformity (0 CFU 100 mL⁻¹), low (1–10), intermediate (11–100), high (101–1000), or very high (> 1000). We reported the distribution of risk categories for each waterpan and season, preserving within-visit variability. We assessed compliance with the requirement that *E. coli* should not be detectable in 100 mL at two levels: the individual sample and the waterpan visit (within-pan positions A, B, and C).

## 3. Results

### 3.1. Spatial and seasonal variation in water quality

Water quality did not differ among the three sampling positions within waterpans (A, human collection point; B, livestock watering point; C, undisturbed area) for any of the 15 parameters (Table S1; F2,210 ≤ 1.58 for the 14 physico-chemical variables, χ²2 = 3.18 for *E. coli*). All 45 pairwise comparisons fell within a pre-specified equivalence margin of 0.5 SD (all p < 0.001), and the largest difference was 0.141 SD, for total phosphorus. *E. coli* at the livestock watering point did not differ from that at the other two positions. In a PCA of all 318 samples (PC1 22.4%, PC2 17.4%), positions overlapped completely (Figure 2a), whereas dry-and wet-season samples separated along PC2 (Figure 2b). The three positions were therefore averaged for each waterpan and occasion, giving 106 waterpan-level observations for physicochemical parameters and 82 for *E. coli*, which were used in all subsequent analyses.

**Fig. 2.**
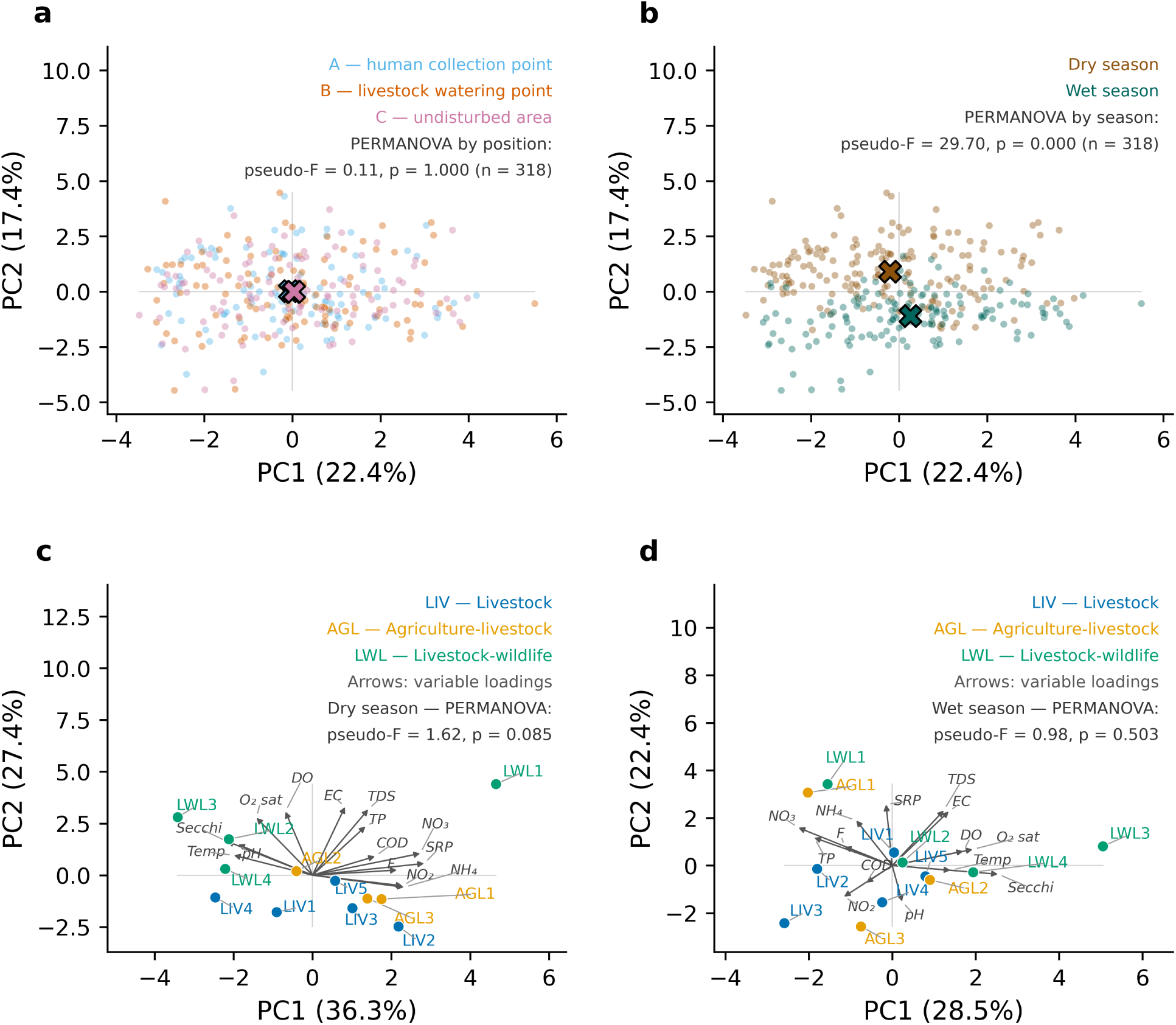
Principal component analysis (PCA) of water quality. (a) All samples (n = 318) coloured by sampling position within the waterpan (A, human collection point; B, livestock watering point; C, undisturbed area). (b) The same ordination coloured by season; large crosses mark group centroids. (c, d) Waterpan means in the (c) dry and (d) wet season (n = 12 per season), coloured by land use (AGL, agriculture–livestock; LIV, livestock; LWL, livestock–wildlife). Arrows show variable loadings at a common scale. All 14 variables were standardised, and skewed variables were log₁₀-transformed in (a, b). Panels a–b and c–d are separate ordinations, so axes are not comparable between them

The first two principal components of the PCAs of seasonal waterpan means explained 63.7% of the variation in the dry season (Figure 2c) and 50.9% in the wet season (Figure 2d). In the dry season, PC1 contrasted clear, warm, alkaline water with nutrient-rich water (SRP, nitrate, ammonium, nitrite) and fluoride, and PC2 reflected ionic content, oxygenation, and TP. LWL1 scored highest on both axes, while LWL2–LWL4 lay on the clear, alkaline side of PC1. In the wet season, PC1 contrasted clear, oxygenated water (LWL3) with water higher in nitrate, TP, nitrite and fluoride, and PC2 was driven by SRP, ionic content and ammonium. AGL1 and LWL1 grouped together with high SRP, nitrate and ammonium. Waterpans did not group by land use in either season, and LWL1 and LWL3 lay on opposite ends of PC1 in both.

Waterpan identity explained more of the variation in water quality composition than season, and land use was not significant (Table S6). Among visits, waterpan explained 20.1% of the variance (F₁₁,₉₄ = 2.15, p < 0.001), although dispersion also differed among waterpans (F = 1.64, p = 0.029; Table S7), so part of this effect may reflect unequal variability within them. Season explained 9.6% after accounting for waterpan (F₁,₁₀₄ = 11.08, p < 0.001), with homogeneous dispersion between seasons. At the waterpan level (n = 12), land use explained 31.8% of the variance but was not significant (F₂,₉ = 2.10, p = 0.051); the season × land-use interaction was also not significant (p = 0.310), and dispersion was homogeneous for both terms (Table S7). Because these terms are tested at the waterpan level, their R² values are not comparable with those for waterpan and season.

Season significantly predicted eight of the 14 water quality parameters, all higher in the dry season: pH (0.65 units higher), TDS, EC, TP, nitrate, nitrite, fluoride and COD (Table 3). The pattern held in every waterpan except for nitrite, which was higher in the wet season in LIV3 and LWL4 (Figure 3). Ammonium tended to be higher in the dry season, but the difference was not significant. Temperature, dissolved oxygen, oxygen saturation, Secchi depth and SRP did not differ between seasons, and neither land use nor waterpan area predicted any parameter (all p > 0.05). Waterpan identity added variation beyond the fixed effects for EC (R²m = 0.375, R²c = 0.628) and Secchi depth (0.215, 0.636), and to a lesser extent for TDS (0.250, 0.376), nitrate (0.214, 0.390) and fluoride (0.244, 0.406), but added little or nothing for pH, dissolved oxygen, oxygen saturation, TP, nitrite and COD (Table 3). Within waterpans, pH, fluoride, nitrite and *E. coli* varied considerably among occasions: nitrite exceeded the guideline mainly on single visits, and *E. coli* peaked in January or February 2026 in nine of the 12 waterpans (Figure S1). Of the 14 parameters, only Secchi depth was associated with waterpan depth and increased with depth (p = 0.012; Table S5).

**Fig. 3.**
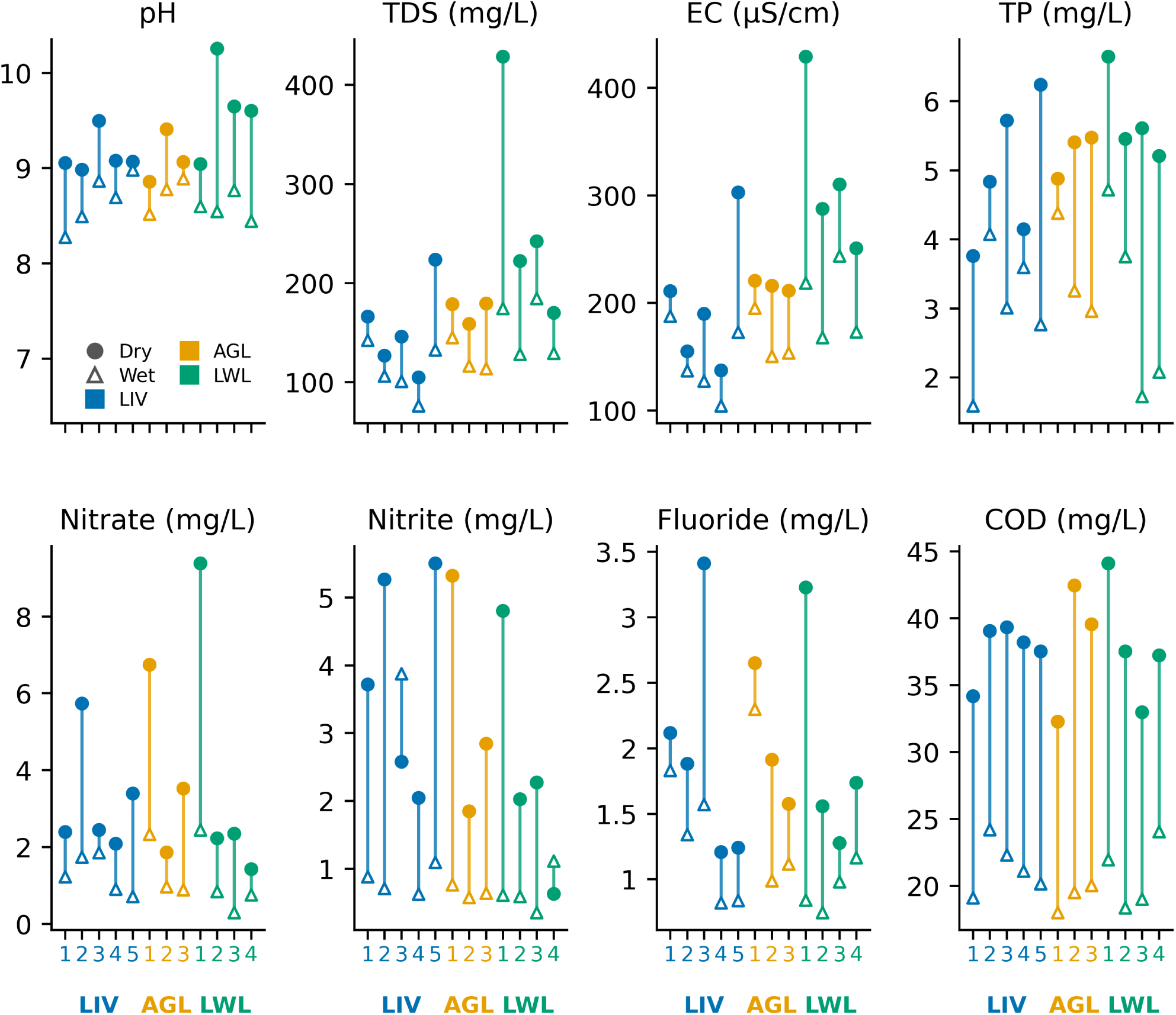
Seasonal mean values of the eight water quality parameters that differed between seasons (Table 3) in each of the 12 waterpans. Filled circles show dry-season and open triangles wet-season means, joined by a vertical line. Waterpans are numbered within land-use groups (LIV, livestock; AGL, agriculture–livestock; LWL, livestock–wildlife). Guideline values (KEBS, 2014; WHO, 2022) are pH 8.5 (WHO upper limit), nitrite 3 mg L⁻¹ and fluoride 1.5 mg L⁻¹

**Table 3.**
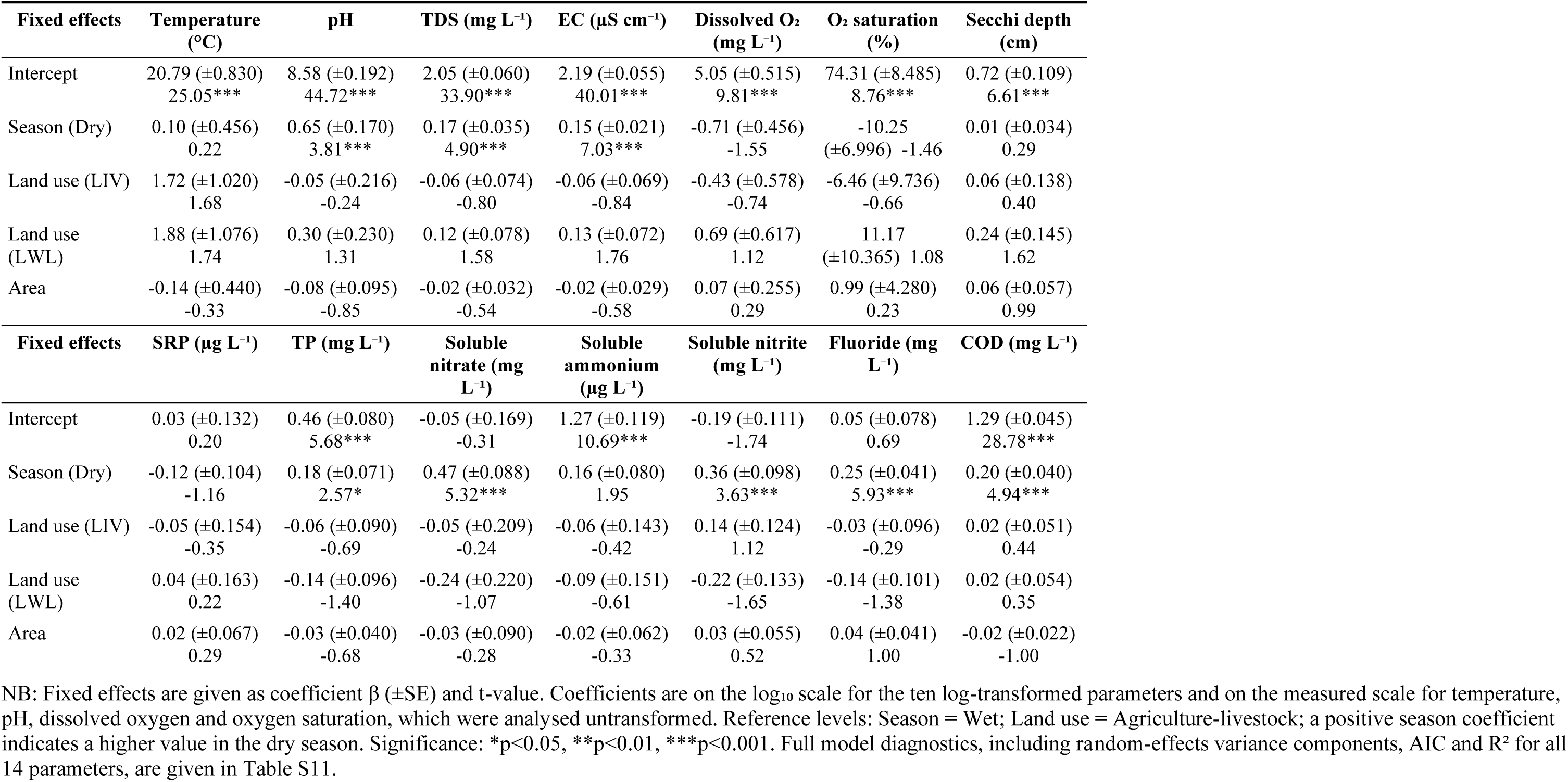
Effects of season, land use, and waterpan area on the 14 physicochemical water quality parameters, from linear mixed-effects models (Equation 2) with waterpan as a random intercept to account for repeated sampling (n = 106 waterpan-occasion observations across 12 waterpans and nine sampling occasions).

### 3.2. Water quality in relation to WHO and KEBS drinking water guidelines

Of the nine parameters with an applicable WHO and/or KEBS guideline, four were above the relevant threshold in at least one waterpan and season: pH, soluble nitrite, fluoride and *Escherichia coli* (Figure 4). TDS, EC, SRP, soluble nitrate and soluble ammonium remained within guideline limits in all waterpans and seasons (Tables S2 and S3). Mean pH was above the WHO upper limit of 8.5 in all 12 waterpans in the dry season and in 9 of the 12 in the wet season; the exceptions were LIV1, LIV2 and LWL4. The wider KEBS limit of 9.5 exceeded only in the dry season, and only in three of the four livestock–wildlife waterpans: LWL2, LWL3 and LWL4, with LWL2 the highest (10.25 ± 1.17).

**Fig. 4.**
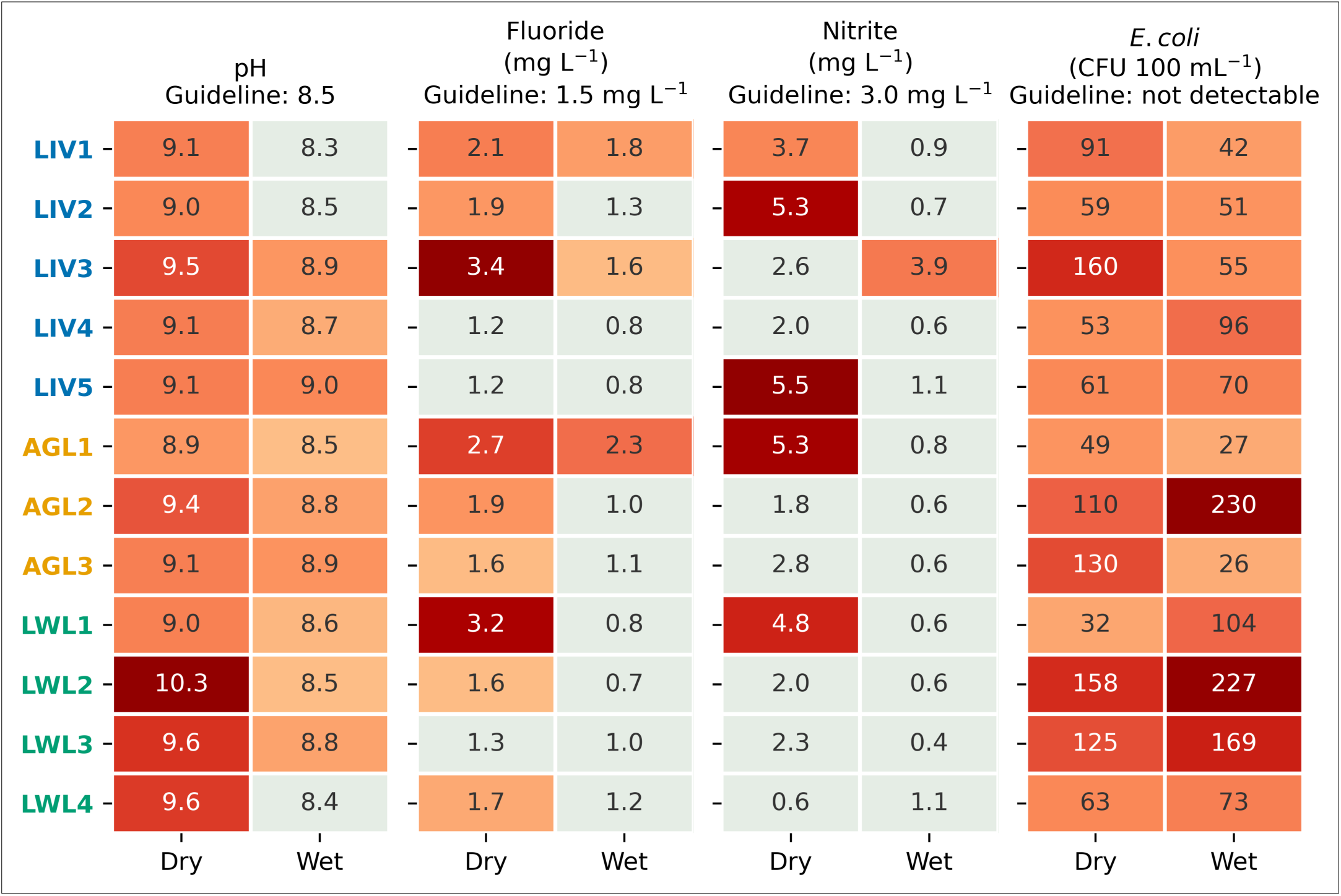
Mean pH, fluoride, nitrite and *E. coli* by waterpan and season, compared with WHO (2022) and KEBS (2014) drinking-water guidelines. Pale cells meet the guideline shown above each panel; orange–red cells exceed it, with darker shades marking larger exceedances. For pH the WHO upper limit of 8.5 is used, and for *E. coli* the guideline is no detectable count, so every cell exceeds it. Waterpan codes are coloured by land use (LIV, livestock; AGL, agriculture–livestock; LWL, livestock–wildlife). Data are from 106 visits (82 for *E. coli*). Other parameters are given in Tables S2 and S3

Fluoride was above the 1.5 mg L⁻¹ guideline in 9 of the 12 waterpans in the dry season, reaching 3.41 ± 1.68 mg L⁻¹ in LIV3, and in only three in the wet season (LIV1, AGL1 and LIV3). Soluble nitrite was above 3 mg L⁻¹ in five waterpans in the dry season (LIV1, AGL1, LIV2, LWL1 and LIV5) (3.72–5.50 mg L⁻¹) and in one in the wet, LIV3 (3.87 ± 6.14 mg L⁻¹). *E. coli* was detected in every waterpan in both seasons, with means from 26 ± 14 CFU 100 mL⁻¹ in AGL3 to 230 ± 267 CFU 100 mL⁻¹ in AGL2, both in the wet season. No waterpan met the requirement that *E. coli* be not detectable in 100 mL. The remaining six parameters (temperature, DO, OS, Secchi depth, TP and COD) have no applicable WHO or KEBS guideline and are reported descriptively (Tables S2 and S3). Overall and season × land-use statistics are given in Table S12.

### 3.3. Effects of season, land use and waterpan area on *Escherichia coli* concentrations

Season and waterpan area did not significantly predict *E. coli* concentrations (Table 4). Land use was also not significant (p = 0.055), although waterpans in livestock–wildlife areas had higher counts than those in agriculture–livestock areas (IRR = 1.71, 95% CI: 1.06–2.75, p = 0.028). Most of the random-effect variation in *E. coli* occurred between sampling occasions (42.8%) and between individual visits (54.7%). Only 2.4% reflected consistent differences between waterpans (Table S8). The full model did not perform better than the null model (likelihood-ratio test, χ²₄ = 4.57, p = 0.335), so season, land use, and waterpan area explained little variation in *E. coli*.

**Table 4.** Negative-binomial mixed model results for *E. coli* counts (n = 246 individual samples, nested within 82 waterpan-occasion combinations across 12 waterpans and seven sampling occasions).

| Fixed effect | Estimate<br>(SE) | z-value | p-value | IRR (95% CI) |
| --- | --- | --- | --- | --- |
| Intercept | 3.833 (0.386) | 9.93 | <0.001*** | 46.19 (21.68–98.41) |
| Season (Dry vs. Wet) | 0.124 (0.463) | 0.27 | 0.788 | 1.13 (0.46–2.81) |
| Land use (Livestock vs. Agriculture-livestock) | 0.069 (0.228) | 0.30 | 0.761 | 1.07 (0.68–1.68) |
| Land use (Livestock-wildlife vs. Agriculture-livestock) | 0.535 (0.243) | 2.20 | 0.028* | 1.71 (1.06–2.75) |
| Waterpan area (standardised) | −0.091<br>(0.102) | −0.90 | 0.371 | 0.91 (0.75–1.11) |

NB: Model: *E. coli* ~ Season + Land use + Area + (1 | Waterpan) + (1 | Occasion) + (1 | Waterpan:Occasion) (Equation 3), fitted by maximum likelihood with glmmTMB (Brooks et al., 2017; family = nbinom1, log link). Reference categories: Season = Wet; Land use = Agriculture-livestock. Waterpan area was standardised (z-score) before fitting. IRR, incidence rate ratio, with 95% Wald confidence intervals. Significance: \* p < 0.05, \*\*\* p < 0.001. Full model diagnostics, including random-effects variance components, dispersion, likelihood-ratio and Wald test statistics, are given in Table S8.

### 3.4. Fluoride exposure risk for children and adults across waterpans and seasons

Mean fluoride concentrations across individual samples were 1.94 mg L⁻¹ in the dry season and 1.21 mg L⁻¹ in the wet. The highest concentration (7.73 mg L⁻¹) was recorded at LWL1 in November 2025, and fluoride varied more at LWL1 than at any other waterpan in either season (Figure 5). Mean child hazard quotients (Equations 4 and 5) exceeded 1 at all 12 waterpans in the dry season and at eight of the 12 in the wet (Figure 5; Table S9). Mean adult quotients exceeded 1 at four waterpans in the dry season (LIV1, AGL1, LIV3 and LWL1) and at one in the wet (AGL1). As expected from the seasonal increase in fluoride (Table 3), hazard quotients were higher in the dry season at every waterpan. At the sample level, 92.0% of dry-season samples and 52.8% of wet-season samples exceeded the child threshold, compared with 27.6% and 14.6% for adults. The highest sample, at LWL1, had a child HQ of 8.58 and an adult HQ of 3.68 (Table S9). Table S10 gives hazard quotients pooled across both seasons.

**Fig. 5.**
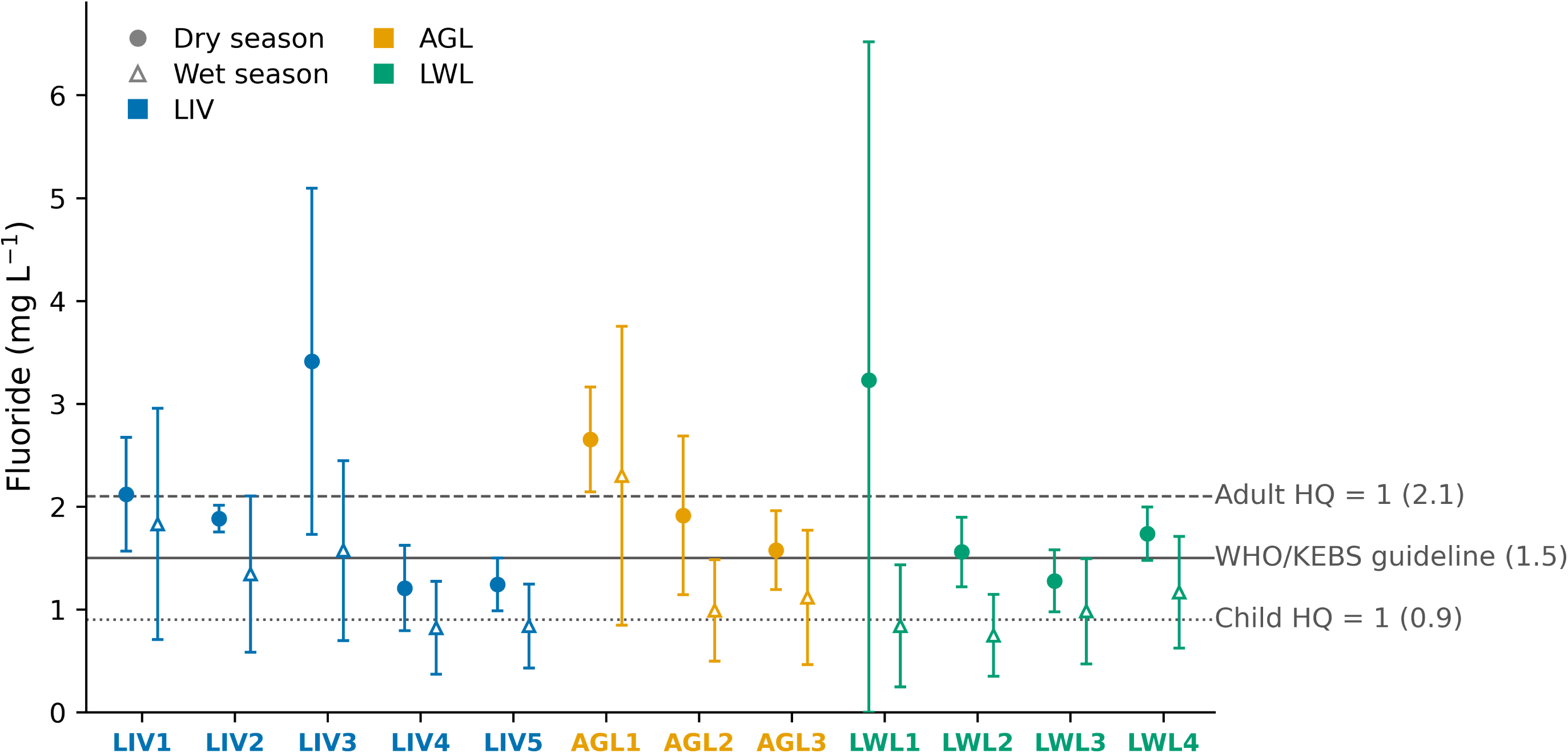
Mean fluoride concentration in each waterpan by season (n = 9–15 samples per waterpan and season). Error bars are ±1 SD, truncated at zero. Horizontal lines mark the WHO/KEBS guideline (1.5 mg L⁻¹) and the concentrations at which the hazard quotient reaches 1 for adults (2.1 mg L⁻¹) and children (0.9 mg L⁻¹). Waterpan codes are coloured by land use (LIV, livestock; AGL, agriculture–livestock; LWL, livestock–wildlife). Hazard quotients by waterpan and season are given in Table S9

### 3.5. Temporal variation in *E. coli* concentrations and the WHO drinking water risk

*E. coli* concentrations varied more between sampling campaigns than between waterpans or seasons (Figure 6; Tables 4 and S8). Between May and December 2025, the median of all samples ranged from 21.0 to 51.5 CFU 100 mL⁻¹ and was lowest in November 2025. It rose to 164.0 CFU 100 mL⁻¹ in January 2026 and was 96.0 CFU 100 mL⁻¹ in February 2026. This increase occurred in all three land-use categories. Median concentrations across all campaigns were 51.0 CFU 100 mL⁻¹ in the dry season and 44.5 CFU 100 mL⁻¹ in the wet season, and season had no significant effect (Table 4).

**Fig 6.**
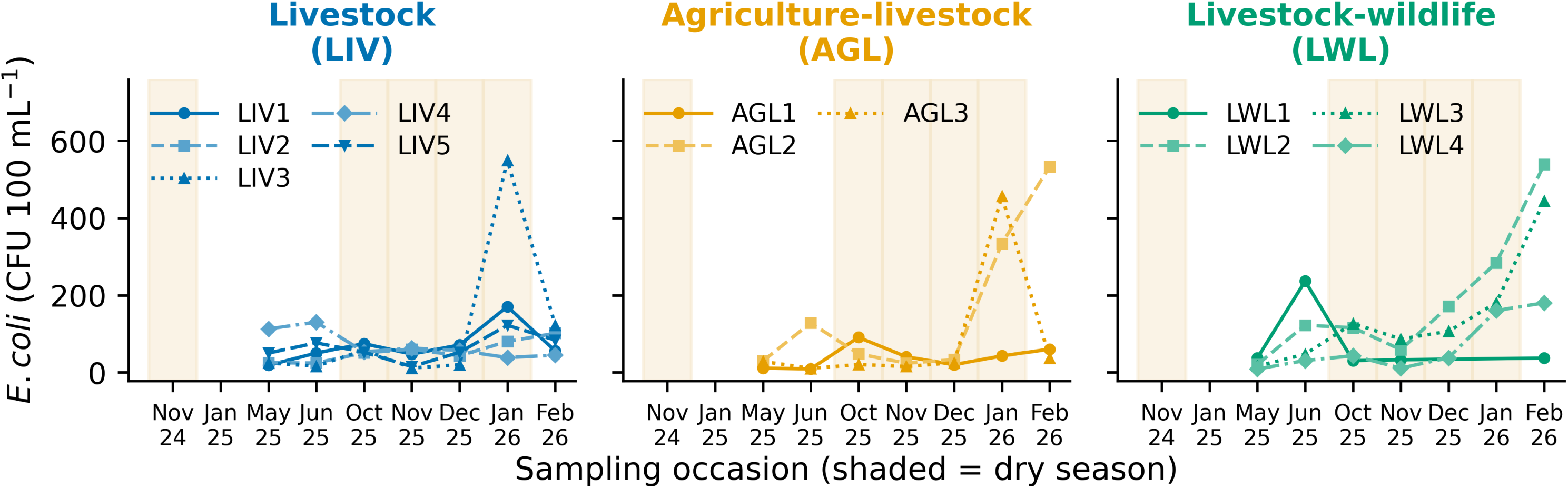
*E. coli* concentration (CFU 100 mL⁻¹) at each waterpan by land-use category. *E. coli* was measured in seven of the nine campaigns (May 2025 to February 2026). Lines join the median of the three sampling points at each visit, and line style and marker identify individual waterpans. Shaded bands mark campaigns classified as dry from household-reported rainfall in the 1–2 weeks before sampling (Section 2.6.1). LWL1 was dry in December 2025 and January 2026

Against WHO (1997) risk categories for drinking water, only one of the 246 individual samples conformed (0 CFU 100 mL⁻¹; AGL3, May 2025). The median for that visit was 23 CFU 100 mL⁻¹, so no waterpan visit met the WHO requirement. An additional 9.3% of samples fell in the low-risk category, 64.2% in the intermediate category, and 26.0% in the high-risk category; none reached the very-high category (> 1000 CFU 100 mL⁻¹; Table S13). The proportion of high-risk samples increased from 5.6% (2 of 36 samples) in May and November 2025 to 66.7% (22 of 33) in January 2026. High-risk samples were most frequent in livestock–wildlife waterpans (31 of 78 samples, 39.7%), against 17.5% in agriculture–livestock and 21.0% in livestock waterpans.

## 4. Discussion

This study reports three major findings. First, microbial contamination consistently compromised drinking-water quality despite little variation among sampling positions within individual waterpans. *Escherichia coli* was detected in all 82 waterpan visits and in 245 of 246 samples, indicating universal microbiological non-compliance, while most physicochemical variables remained within guideline values. Fluoride and the combined nitrate–nitrite criterion were the principal chemical concerns, with exceedances concentrated particularly during the dry season. Second, microbial and chemical hazards differed in their temporal and spatial behaviour. Fluoride concentrations were consistent with the regional geochemical setting and tended to increase during drawdown periods, whereas *E. coli* varied predominantly among sampling occasions and individual visits, with little evidence of persistent differences among waterpans or broad land-use classes. Third, we could not reduce water-quality variation to a single dominant environmental driver. Individual waterpans differed substantially in multivariate composition, while campaign-level dry–wet effects were strongest for electrical conductivity and inorganic nitrogen but became less certain after accounting for the limited number of independent campaigns. Land use poorly predicted water quality and microbial contamination. Together, these findings point to heterogeneous, multi-hazard risks requiring both routine microbial protection and targeted, waterpan-specific chemical monitoring.

### 4.1. Factors influencing water quality: seasonal and waterpan-specific controls

Season had the greatest influence on water quality of the three factors tested. All eight parameters that differed between seasons were higher in the dry season, in every waterpan except for nitrite in LIV3 and LWL4 (Table 3; Figure 3). TDS and EC were about 48% and 41% higher, consistent with evaporative concentration as volume contracts and residence time lengthens, and the contrast was greatest in LWL1 (Figure 3). Where rainfall dominates the water balance, conductivity instead falls with wet-season dilution (Kotut et al., 1999; Lencha et al., 2021). The higher dry-season pH (0.65 units) and oxygen saturation up to 188% (Table S12) suggest net photosynthetic production in shallow water, as reported for Turkwel Gorge Reservoir (Kotut et al., 1999) and Nadosoito Dam (Eliakimu et al., 2018). All samples were taken in daylight, so these values describe daytime conditions. High pH may also explain why TP rose while SRP did not, since high pH releases phosphorus from sediments, which is then rapidly taken up (Eliakimu et al., 2018; Woldeab et al., 2018). Nitrite is an intermediate of nitrification, and its accumulation may reflect a temperature-dependent imbalance between ammonia and nitrite oxidation (Taylor & Mellbye, 2022). Higher COD is consistent with organic matter accumulating where inflow is negligible (Benariba et al., 2022). Temperature, dissolved oxygen, oxygen saturation and Secchi depth did not differ between seasons. Shallow depth and wind mixing probably keep conditions uniform and prevent the dry-season oxygen decline reported for deeper impoundments such as Kiri Reservoir (Hassan, 2026). Season did not predict *E. coli* (Table 4), which fits a supply dominated by direct animal deposition rather than runoff (Yakirevich et al., 2026). We did not sample runoff, so we cannot exclude a first-flush input at the onset of the rains.

Land use did not predict any of the 14 parameters, and waterpans did not group by land use in either season (Table 3; Figure 2). Land use and area are waterpan-level covariates with 12 replicates, so their tests have less power than the season term. Livestock also use all three categories, narrowing the contrast in faecal and organic inputs the classification was meant to capture. Land use alone likewise failed to explain farm-dam water quality (Brainwood et al., 2004), and pond-specific characteristics explained much of the nutrient variation across 240 ponds (Bartrons et al., 2025). Land use was marginally associated with *E. coli*, with higher counts in livestock–wildlife than agriculture–livestock catchments (Table 4).

Waterpan identity explained about twice as much of the variation in composition as season (Table S6), and dispersion also differed among pans (Table S7). The measured waterpan characteristics explained little of this: depth predicted only Secchi depth, and area predicted no parameter (Tables 3, 4, and S5). Among-pan differences were strongest for EC, Secchi depth, TDS, nitrate and fluoride, constituents tied to the setting of each pan rather than to processes within the water column (Table 3). Geology is the most likely unmeasured attribute. The pans lie on Cenozoic phonolites, trachytes, and basalts interbedded with volcanic ash and tuff, with Precambrian quartzites to the south-west (Saggerson, 1966; Wright, 1967); saline springs in the area are attributed to sodium salts leached from pyroclastics (Olang, 2025; Tuna, 2025). Spatially variable elevated fluoride is consistent with this setting. In the southern Kenya Rift, alkaline volcanic rocks, alkaline groundwater and prolonged water–rock interaction, promote fluoride mobilisation (Gaciri & Davies, 1993; Olaka et al., 2016). Hydrochemical and isotopic work across the wider Narok landscape shows that pans differ in their connection to groundwater and springs, with fluoride loading, along with bicarbonate, sodium, and manganese, in groundwater-influenced pans (Tuna, 2025). The pH of pans in this landscape ranged from 7.1 to 10.0 (Olang, 2025). At high pH, in sodium–bicarbonate water with little calcium, fluorite precipitation is unlikely to limit fluoride, so fluoride can persist and rise further as waterpans draw down, which fits the higher dry-season concentrations (Table 3). This study did not quantify groundwater inputs, mineralogy or source-specific tracers. The fluoride pattern is therefore consistent with regional geogenic control, but it does not directly show that fluoride in individual pans is geogenic.

### 4.2. Drinking water quality and compliance

Water quality did not differ among the three sampling positions (human collection point, livestock watering point, and undisturbed point) for any of the 15 parameters, and all 45 pairwise comparisons fell within the 0.5 SD equivalence margin (Figure 2a, Table S1). This uniformity suggests rapid mixing in the pans. Their shallow depth and intensive use by humans, livestock and wildlife disturb the water column frequently enough to mix inputs faster than they can accumulate at any one point. This is similar to findings at Kiri Reservoir, which is well mixed and where water quality varied little among locations (Hassan, 2026). Oyhakilome et al. (2012) likewise reported that seasonal differences were significant at Owena Dam, while differences among sampling points were not. These findings contrast with those from deeper, less disturbed small waterbodies (Pachepsky et al., 2018; Rashedi et al., 2026). In Maryland irrigation ponds, Pachepsky et al. (2018) reported temporarily stable within-pond patterns producing ten-fold differences in *E. coli* between locations. Those ponds had surface areas similar to the waterpans studied here (about 0.40 ha vs a mean of 0.45 ha) but were much deeper, averaging 2.7 m, and were not used by livestock. Livestock access reduces this spatial structure: across 32 ponds, geochemistry explained 24% of microbial community variation in undisturbed ponds versus 10% in cattle-accessed ponds (Zahir et al., 2026), but only between ponds, not within them.

One major finding of this study is that *E. coli* was detected in every waterpan in both seasons (Figure 4), and in 245 of the 246 samples. The single negative sample (AGL3, May 2025) came from a visit where the other two samples were positive, so that visit failed the WHO and KEBS requirement that *E. coli* be undetectable in 100 mL of drinking water. Because *E. coli* indicates recent faecal contamination by warm-blooded animals, contamination was widespread rather than confined to particular pans. The WHO/UNICEF Joint Monitoring Programme (JMP) classifies surface water as the lowest drinking-water service level, below other unimproved sources (WHO/UNICEF, 2023). Open surface waters elsewhere in Kenyan drylands show the same pattern. *E. coli* occurred in 62.5% of samples from shared surface waterpoints in Narok County (Waswala-Olewe et al., 2026), and in Isiolo County only 36.8% of surface-water samples were free of *E. coli*, *Staphylococcus aureus* and *Clostridium perfringens*, compared with 77.1% of groundwater samples free of *E. coli* alone (Onyango et al., 2018). Animal droppings and urine were identified as possible pollutant sources in an earthen dam serving pastoral communities in northern Tanzania (Eliakimu et al., 2018).

Chemical risk was much lower. TDS, EC, SRP, nitrate and ammonium stayed within guidelines in every waterpan and season, usually by wide margins (Tables S2 and S3). TDS and EC peaked in LWL1 in the dry season (428 mg L⁻¹ and 429 µS cm⁻¹), still below the guideline, and nitrate remained far below the WHO (50 mg L⁻¹) and KEBS (45 mg L⁻¹) guidelines (waterpan means ≤ 9.4 mg L⁻¹). The pans are therefore not generally mineralised or nutrient-polluted. The chemical risk lies in alkalinity, fluoride and, locally, nitrite, whereas microbial contamination was the dominant hazard. A similar contrast was reported for the Owena Dam in Nigeria, where most parameters were acceptable but microbial load, turbidity, colour, iron and manganese required treatment before supply (Oyhakilome et al., 2012).

Mean pH was 9.01. It exceeded the WHO limit of 8.5 in every waterpan in the dry season and in nine of 12 in the wet season, reaching 10.25 in LWL2. Only three livestock–wildlife pans exceeded the KEBS limit of 9.5 in the dry season (Figure 4; Table S12). Comparable values are reported for surface waterpoints and waterpans elsewhere in Narok County (Moseti et al., 2025; Waswala-Olewe et al., 2026), so this appears to be a property of shallow alkaline impoundments, not of pollution. It does matter for treatment. Boiling and chlorination were the most commonly reported household treatments, and chlorine disinfects effectively only at pH below about 8 because, above this, hypochlorous acid dissociates to the weakly germicidal hypochlorite ion (WHO, 2022). Chlorination at these pH values would be less effective unless the dose or pH is adjusted, whereas boiling is unaffected. Fluoride exceeded the 1.5 mg L⁻¹ guideline in nine of 12 waterpans in the dry season (up to 3.41 mg L⁻¹ in LIV3) and in three in the wet season (LIV1, LIV3 and AGL1; Figure 4), a pattern consistent with the volcanic setting discussed in Section 4.1. Nitrite exceeded 3 mg L⁻¹ in five waterpans in the dry season (up to 5.50 mg L⁻¹ in LIV5) and in LIV3 in the wet season. The nitrite guideline protects bottle-fed infants from methaemoglobinaemia, and WHO notes that gastrointestinal infection raises this risk (WHO, 2022). Both conditions are present here, so the nitrite exceedances matter despite low nitrate.

### 4.3. Fluoride exposure and health risk

The fluoride hazard was greater for children than for adults. Mean child hazard quotients exceeded 1 at every waterpan in the dry season, while adult quotients did so at only four (LIV1, LIV3, AGL1 and LWL1; Figure 5; Table S9). This difference reflects intake per unit body weight. A child drinking 1 L day⁻¹ at 15 kg takes in more than twice as much fluoride per kilogram as an adult drinking 2 L day⁻¹ at 70 kg (Table 2), so a quotient of 1 is reached at 0.9 mg L⁻¹ for a child and 2.1 mg L⁻¹ for an adult (Equation 5; Figure 5). Water at the 1.5 mg L⁻¹ guideline therefore carries a quotient of 1.67 for a child and 0.71 for an adult, and compliance with the guideline does not protect children. Fluoride risk assessments elsewhere report the same age gradient (Guissouma et al., 2017; Yousefi et al., 2018).

Dry-season mean concentrations exceeded 2 mg L⁻¹ in four of the 12 waterpans (Table S9). In a Kenyan population drinking water at about this concentration, all 102 children examined had dental fluorosis (Manji et al., 1986), so the concentrations in our study may pose a risk of dental fluorosis. Fluoride did not vary with land use but differed consistently among pans (Table 3), suggesting a geogenic source (Section 4.1), although we did not measure source-specific tracers. Concentrations in the pans are modest compared with other Rift Valley waters, where Lake Nakuru has been reported at up to 2,800 mg L⁻¹ (Malago et al., 2017), but the pans are small standing waters on which dryland communities depend for drinking. Fluoride rose in the dry season alongside TDS and EC (Table 3), consistent with evaporative concentration as volume declined. LWL1 had the highest exposure of the 12 pans (Table S9) and dried out later in the same season.

Peak exposure coincides with peak dependence, because fluoride is highest in the dry season, when reliance on waterpans is greatest. Dry-season concentrations, not annual means, therefore describe the greatest exposure. Concentrations also varied widely within a pan: in LWL1, visit means ranged from 0.45 to 6.99 mg L⁻¹. A single measurement can therefore misjudge the risk at an individual waterpan.

### 4.4. Faecal contamination: temporal variation and sources

*E. coli* varied mainly by sampling occasion and visit, and neither season nor waterpan area was significant (Tables 4 and S8). Contamination was therefore episodic, whereas fluoride varied with waterpan and season (Figure 5). Counts peaked in January–February 2026 in all three land-use categories (Figure 6). Between peaks, mean counts at every waterpan exceeded the low-risk category in both seasons, and no sample reached the very-high category (Table S13), so contamination was widespread and moderate in magnitude. Under the WHO (1997) classification, waterpan–season means fell in the intermediate and high-risk categories, and *E. coli* in drinking water is associated with diarrhoeal disease in household settings (Gruber et al., 2014). Conditions in the pans may also favour persistence. Sunlight inactivation of *E. coli* declines with water absorbance and depth (Maraccini et al., 2016), and at a mean Secchi depth of 8.30 cm (Table S12) light penetrates only the upper centimetres, leaving most of the water column shielded. Temperature did not differ between seasons (Table 3; Table S2), so seasonal differences in thermal die-off are unlikely.

Contamination may also be sustained within the pans. Livestock use them daily and disturb the bottom sediments. In tropical lakes, *E. coli* moves from sediment into the water column faster than it decays, which can maintain counts without recent faecal input (Liu et al., 2024), and in tropical soils *E. coli* has multiplied and remained metabolically active in response to nutrient inputs (Byappanahalli et al., 2012). Some counts may therefore reflect resident populations within the pans and not only recent deposition. Sediment was not sampled, so this remains an inference.

These conditions were common to all 12 pans, and differences among them were small relative to variation between visits. Nevertheless, high-risk samples were about twice as frequent in livestock–wildlife catchments as in the other categories (IRR = 1.71, p = 0.028; Table 4). Land use overall was not significant (χ²₂ = 5.80, p = 0.055), and the full model fitted no better than one without fixed effects (ΔAIC = +3.43; Table S8), so the effect is suggestive, not established. Because land use is a waterpan-level term with 12 replicates (Section 4.1), the test is better read as inconclusive than as evidence against an effect. Joint use by livestock and wildlife may contribute, as in a reservoir where cattle and migratory waterfowl each contributed significantly to *E. coli* concentrations (Hansen et al., 2020), but this was not tested here. Livestock, wildlife and humans all use these pans, and apportioning the load among them requires microbial source tracking.

## 5. Conclusions

Waterpans in Narok County pose two drinking-water hazards driven by different factors. *E. coli* was detected at every waterpan visit and varied between sampling occasions, not with season or sampling position, so faecal contamination was episodic but always present. Chemical quality met the guidelines except for pH, nitrite and fluoride, which exceeded them at more waterpans in the dry season, when most constituents were higher. Waterpan identity explained about twice as much variation in composition as season, but not across the area or depth we measured; land use was inconclusive with only 12 waterpans. Fluoride put children above the hazard threshold at every waterpan in the dry season, so compliance with the 1.5 mg L⁻¹ guideline does not protect them. The differences in fluoride among pans are consistent with geogenic control, which this study did not test directly.

Management therefore needs two tracks. Faecal risk calls for protection in every season: protected abstraction points separated from livestock watering, and point-of-use treatment for the residual exposure, because wildlife access cannot readily be controlled. Boiling is unaffected by the alkaline pH (mean 9.0), whereas chlorination needs dose adjustment, and neither removes fluoride or nitrite. Fluoride requires either fluoride-specific treatment or reduced reliance on the pans during the dry season, through wet-season rainwater harvesting or access to a perennial supply, and water above the nitrite guideline should not be used to prepare infant formula. For monitoring, one sampling position per pan was representative, so effort is better spent on more waterpans, with repeated visits where faecal contamination is the concern. Microbial source tracking and sediment sampling are needed to resolve wildlife input and sediment resupply of *E. coli*, and groundwater and mineralogical tracers to establish the fluoride source.

## Statements and Declarations

### Author contributions

Sharon F. Akaranga: Conceptualization, Methodology, Investigation, Data curation, Formal analysis, Visualization, Writing – original draft, Writing – review & editing. Edith J. Kurui: Investigation, Data curation, Writing – review & editing. Kelvin O. Moenga: Investigation, Data curation, Formal analysis, Writing – review & editing. Claire Furlong: Conceptualization, Methodology, Investigation, Funding acquisition, Supervision, Writing – review & editing. Jack van de Vossenberg: Conceptualization, Methodology, Investigation, Supervision, Writing – review & editing. Frank O. Masese: Conceptualization, Methodology, Investigation, Funding acquisition, Project administration, Formal analysis, Visualization, Supervision, Writing – review & editing.

### Ethics approval

Ethical approval was granted by the Institutional Research and Ethics Committee of Moi University/Moi Teaching and Referral Hospital (IREC; Reference IREC/1257/2025; Approval No. 0005198), and a research licence was issued by the National Commission for Science, Technology and Innovation (NACOSTI; License No. NACOSTI/P/25/4181282). County research authorisation was granted by the County Director of Education, Narok County (Ref. CDE/NRK/RES/VOL.11/43). Permission to access and sample the waterpans was obtained from local community leaders and waterpan committees, area chiefs and the local administration, and, where waterpans were on private land, from the landowners. All procedures involving human participants were conducted in accordance with the ethical standards of the IREC.

### Consent to participate

Informed consent was obtained from all household respondents who provided information used in this study.

### Consent to publish

Not applicable. This article contains no data, images or other information that could identify any individual participant.

### Data availability

Data will be made available on request.

## Acknowledgements

We thank the households and community leaders of the twelve waterpan communities along the Mulot–Narok and Narok–Sekenani transects, Narok County, for access to their waterpans and for generously sharing their time and knowledge. We thank the wider Sustainable Waterpans Project team at the University of Eldoret and IHE Delft Institute for Water Education. We thank Christian Bashizi for help with mapping the study area, and Elizabeth Wanderi for assistance with field sampling. We are grateful to Henry Lubanga and Mary Kiplagat (Department of Fisheries and Aquatic Sciences) and Margaret Maina and Edwin Kirwa (Department of Chemistry), University of Eldoret, for support with laboratory analyses, and to the Department of Fisheries and Aquatic Sciences, University of Eldoret, for hosting the study and providing field equipment and laboratory facilities.

