## Supplementary Material for "Microbial and chemical hazards in multi-use dryland waterpans: temporal variability, local heterogeneity and implications for drinking-water safety"

*Journal: Environmental Geochemistry and Health*

### Contents

Table S1. Comparison of water quality among the three within-waterpan sampling positions
Table S2. Physicochemical water quality by waterpan and season, with WHO and KEBS guideline values
Table S3. Nutrients, fluoride, organic matter and *E. coli* by waterpan and season, with guideline values
Table S4. Sampling campaigns, dates, season and number of visits
Table S5. Effect of waterpan depth on the 14 physicochemical parameters
Table S6. PERMANOVA of water quality composition
Table S7. Homogeneity of multivariate dispersion
Table S8. Full negative-binomial mixed model results for *E. coli*
Table S9. Fluoride exposure and hazard quotients by waterpan and season
Table S10. Fluoride exposure and hazard quotients by waterpan, pooled across seasons
Table S11. Full linear mixed-effects model results for the 14 physicochemical parameters
Table S12. Water quality across all waterpans and sampling occasions, by season and land use
Table S13. *E. coli* concentrations as WHO risk categories, by waterpan and season
Fig. S1 Time series of pH, fluoride, nitrite and *E. coli* in each waterpan

**Table S1.** Tests of physicochemical and microbial differences among the three within-waterpan sampling positions (human collection point, livestock watering point and comparatively undisturbed area). LMM, linear mixed model (Equation 1 in the main text), with ($\log_{10}$) indicating a $\log_{10}$-transformed response; NB GLMM, negative-binomial generalised linear mixed model; p (FDR), Benjamini–Hochberg adjusted p value across the 15 parameters; Largest difference (SD), largest pairwise difference between positions, in standard deviation units.

| Parameter (unit) | Model | n | df | Statistic | p | p (FDR) | Largest difference (SD) |
|---|---|---|---|---|---|---|---|
| Temperature (°C) | LMM | 318 | 2, 210 | F = 0.358 | 0.699 | 0.971 | 0.054 |
| pH | LMM | 318 | 2, 210 | F = 0.302 | 0.739 | 0.971 | 0.037 |
| Dissolved oxygen (mg $L^{-1}$) | LMM | 318 | 2, 210 | F = 0.856 | 0.426 | 0.971 | 0.070 |
| Oxygen saturation (%) | LMM | 318 | 2, 210 | F = 1.294 | 0.276 | 0.971 | 0.094 |
| TDS (mg $L^{-1}$) | LMM ($\log_{10}$) | 318 | 2, 210 | F = 0.261 | 0.770 | 0.971 | 0.018 |
| EC (µS $cm^{-1}$) | LMM ($\log_{10}$) | 318 | 2, 210 | F = 0.274 | 0.761 | 0.971 | 0.024 |
| SRP (µg $L^{-1}$) | LMM ($\log_{10}$) | 318 | 2, 210 | F = 0.098 | 0.906 | 0.971 | 0.025 |
| TP (mg $L^{-1}$) | LMM ($\log_{10}$) | 318 | 2, 210 | F = 1.575 | 0.210 | 0.971 | 0.141 |
| Soluble $NO_3^-$ (mg $L^{-1}$) | LMM ($\log_{10}$) | 318 | 2, 210 | F = 0.308 | 0.735 | 0.971 | 0.048 |
| Soluble $NH_4^+$ (µg $L^{-1}$) | LMM ($\log_{10}$) | 318 | 2, 210 | F = 0.331 | 0.719 | 0.971 | 0.046 |
| Soluble $NO_2^-$ (mg $L^{-1}$) | LMM ($\log_{10}$) | 318 | 2, 210 | F = 0.012 | 0.988 | 0.988 | 0.010 |
| Fluoride (mg $L^{-1}$) | LMM ($\log_{10}$) | 318 | 2, 210 | F = 1.185 | 0.308 | 0.971 | 0.056 |
| COD (mg $L^{-1}$) | LMM ($\log_{10}$) | 318 | 2, 210 | F = 0.223 | 0.800 | 0.971 | 0.031 |
| Secchi depth (cm) | LMM ($\log_{10}$) | 318 | 2, 210 | F = 0.107 | 0.899 | 0.971 | 0.021 |
| *E. coli* (CFU 100 $mL^{-1}$) | NB GLMM | 246 | 2 | $\chi^2$ = 3.178 | 0.204 | 0.971 | 0.093 |

NB: No parameter differed among positions, and *E. coli* was not higher at the livestock watering point; the three positions were therefore averaged for each waterpan and sampling occasion (main text, Section 3.1).

**Table S2.** Physicochemical water quality parameters by waterpan and season, compared with WHO (2022) and KEBS (2014) drinking water guideline values.

| Waterpan (Land use) | Season | n | Temperature (°C) | pH | TDS (mg $L^{-1}$) | EC (µS $cm^{-1}$) | Dissolved $O_2$ (mg $L^{-1}$) | $O_2$ saturation (%) | Secchi depth (cm) |
|---|---|---|---|---|---|---|---|---|---|
| *Guideline (WHO / KEBS)* | | | – | *6.5–8.5 / 5.5–9.5* | *1000 / 1500* | *– / 2500* | – | – | – |
| LIV1 (LIV) | Dry | 5 | 23.30 ± 0.75 | **9.06 ± 0.93** | 166.13 ± 54.10 | 211.09 ± 26.07 | 3.70 ± 1.97 | 55.21 ± 29.62 | 10.04 ± 1.84 |
| | Wet | 4 | 22.38 ± 2.67 | 8.27 ± 0.91 | 142.25 ± 41.67 | 187.63 ± 14.91 | 3.95 ± 2.33 | 62.17 ± 39.57 | 8.71 ± 2.39 |
| AGL1 (AGL) | Dry | 5 | 19.98 ± 0.74 | **8.86 ± 0.82** | 178.73 ± 57.32 | 220.63 ± 24.19 | 4.22 ± 2.44 | 60.02 ± 33.18 | 6.44 ± 1.64 |
| | Wet | 4 | 20.84 ± 1.46 | **8.52 ± 0.85** | 145.08 ± 44.45 | 194.97 ± 29.85 | 5.26 ± 2.11 | 75.56 ± 31.03 | 4.15 ± 0.55 |
| LIV2 (LIV) | Dry | 5 | 21.19 ± 1.38 | **8.98 ± 0.64** | 126.71 ± 40.17 | 154.96 ± 14.06 | 3.16 ± 1.92 | 45.38 ± 27.71 | 4.06 ± 0.71 |
| | Wet | 4 | 21.25 ± 2.45 | 8.49 ± 1.12 | 106.25 ± 54.18 | 136.88 ± 33.19 | 5.06 ± 3.13 | 73.71 ± 47.48 | 4.95 ± 1.61 |
| AGL2 (AGL) | Dry | 5 | 23.04 ± 4.71 | **9.41 ± 0.51** | 158.93 ± 50.43 | 215.95 ± 67.58 | 5.17 ± 4.14 | 79.44 ± 65.10 | 3.62 ± 0.79 |
| | Wet | 4 | 22.04 ± 2.85 | **8.78 ± 0.87** | 116.50 ± 65.75 | 150.22 ± 47.50 | 5.70 ± 3.51 | 88.59 ± 54.51 | 7.68 ± 3.35 |
| LIV3 (LIV) | Dry | 5 | 21.55 ± 1.68 | **9.50 ± 0.77** | 145.93 ± 35.93 | 189.73 ± 28.76 | 3.06 ± 1.52 | 44.00 ± 19.03 | 4.13 ± 0.80 |
| | Wet | 4 | 22.76 ± 3.49 | **8.86 ± 1.07** | 100.75 ± 10.11 | 127.37 ± 25.86 | 4.32 ± 1.77 | 58.32 ± 17.32 | 5.11 ± 2.22 |
| AGL3 (AGL) | Dry | 5 | 18.52 ± 2.42 | **9.06 ± 0.73** | 179.53 ± 91.01 | 211.25 ± 59.26 | 3.84 ± 2.08 | 54.13 ± 38.04 | 4.78 ± 0.37 |
| | Wet | 4 | 20.89 ± 3.38 | **8.89 ± 1.13** | 113.52 ± 50.57 | 153.30 ± 36.90 | 3.94 ± 2.70 | 57.20 ± 41.85 | 8.35 ± 0.94 |
| LWL1 (LWL) | Dry | 3 | 21.53 ± 0.43 | **9.04 ± 0.93** | 428.44 ± 116.30 | 428.89 ± 116.44 | 6.08 ± 1.14 | 79.19 ± 24.15 | 5.04 ± 0.04 |
| | Wet | 4 | 21.85 ± 2.73 | **8.60 ± 1.19** | 174.25 ± 114.14 | 218.50 ± 101.74 | 4.94 ± 1.85 | 72.21 ± 26.34 | 4.11 ± 0.72 |
| LIV4 (LIV) | Dry | 5 | 24.89 ± 1.76 | **9.08 ± 0.66** | 105.00 ± 26.31 | 137.20 ± 20.51 | 5.16 ± 2.10 | 77.87 ± 32.21 | 5.77 ± 0.73 |
| | Wet | 4 | 21.69 ± 2.43 | **8.69 ± 1.20** | 76.31 ± 23.41 | 104.42 ± 8.32 | 5.99 ± 3.80 | 86.41 ± 58.76 | 6.15 ± 1.27 |
| LIV5 (LIV) | Dry | 5 | 23.43 ± 1.98 | **9.07 ± 0.60** | 223.73 ± 50.92 | 302.80 ± 57.20 | 3.32 ± 1.91 | 50.27 ± 29.27 | 5.25 ± 0.96 |
| | Wet | 4 | 23.53 ± 2.25 | **8.98 ± 0.78** | 132.43 ± 45.62 | 172.56 ± 22.75 | 4.89 ± 1.54 | 72.56 ± 25.02 | 7.47 ± 1.58 |
| LWL2 (LWL) | Dry | 5 | 23.66 ± 3.30 | **10.25 ± 1.17** | 222.33 ± 54.25 | 287.53 ± 35.76 | 5.20 ± 1.23 | 72.64 ± 12.58 | 11.46 ± 8.28 |
| | Wet | 4 | 23.21 ± 1.78 | **8.54 ± 0.92** | 128.08 ± 56.56 | 167.89 ± 56.42 | 4.66 ± 1.74 | 69.54 ± 25.63 | 6.02 ± 1.95 |
| LWL3 (LWL) | Dry | 5 | 24.63 ± 1.66 | **9.65 ± 1.23** | 242.27 ± 85.72 | 310.20 ± 16.25 | 5.53 ± 2.89 | 98.45 ± 47.07 | 26.31 ± 7.52 |
| | Wet | 4 | 23.68 ± 1.54 | **8.77 ± 0.89** | 184.50 ± 54.35 | 243.67 ± 20.88 | 6.44 ± 3.28 | 95.82 ± 49.39 | 17.22 ± 8.64 |
| LWL4 (LWL) | Dry | 5 | 20.95 ± 1.17 | **9.60 ± 1.02** | 170.00 ± 48.79 | 250.87 ± 26.13 | 4.65 ± 2.47 | 65.37 ± 33.94 | 15.87 ± 5.97 |
| | Wet | 4 | 21.27 ± 1.62 | 8.44 ± 0.40 | 129.33 ± 52.96 | 172.95 ± 21.67 | 6.15 ± 2.29 | 93.28 ± 37.35 | 14.18 ± 9.24 |

NB: Values are mean ± SD of waterpan-visit observations. n, number of sampling visits (total 106). LWL1 was dry in December 2025 and January 2026. A dash indicates no applicable WHO or KEBS guideline. Bold pH values are above the WHO upper limit of 8.5; no other parameter in this table was above its guideline. TDS, total dissolved solids; EC, electrical conductivity. Land use: AGL, agriculture–livestock; LIV, livestock; LWL, livestock–wildlife.

**Table S3.** Nutrient, fluoride, organic matter and *Escherichia coli* concentrations by waterpan and season, compared with WHO (2022) and KEBS (2014) drinking water guideline values.

| Waterpan (Land use) | Season | n (nutrients, $F^-$, COD / *E. coli*) | SRP (µg $L^{-1}$) | TP (mg $L^{-1}$) | Soluble nitrate (mg $L^{-1}$) | Soluble ammonium (µg $L^{-1}$) | Soluble nitrite (mg $L^{-1}$) | Fluoride (mg $L^{-1}$) | COD (mg $L^{-1}$) | *E. coli* (CFU 100 $mL^{-1}$) |
|---|---|---|---|---|---|---|---|---|---|---|
| *Guideline (WHO / KEBS)* | | | *– / 2200* | *–* | *50 / 45* | *– / 500* | *3 / 3* | *1.5 / 1.5* | *–* | *Not detectable in 100 mL* |
| LIV1 (LIV) | Dry | 5 / 4 | 0.97 ± 0.72 | 3.76 ± 3.23 | 2.39 ± 1.07 | 27.65 ± 24.03 | **3.72 ± 3.66** | **2.12 ± 0.55** | 34.18 ± 19.85 | **91 ± 54** |
| | Wet | 4 / 3 | 2.74 ± 3.98 | 1.59 ± 0.85 | 1.23 ± 0.91 | 11.21 ± 11.51 | 0.88 ± 0.56 | **1.83 ± 1.12** | 19.09 ± 2.76 | **42 ± 20** |
| AGL1 (AGL) | Dry | 5 / 4 | 1.38 ± 0.59 | 4.88 ± 3.14 | 6.74 ± 3.89 | 34.41 ± 20.66 | **5.32 ± 3.98** | **2.65 ± 0.51** | 32.27 ± 24.63 | **49 ± 30** |
| | Wet | 4 / 3 | 3.00 ± 3.41 | 4.38 ± 2.61 | 2.33 ± 3.08 | 43.38 ± 13.42 | 0.76 ± 0.26 | **2.30 ± 1.45** | 18.00 ± 1.51 | **27 ± 29** |
| LIV2 (LIV) | Dry | 5 / 4 | 1.69 ± 0.71 | 4.83 ± 3.04 | 5.73 ± 3.41 | 45.36 ± 25.39 | **5.26 ± 4.46** | **1.88 ± 0.13** | 39.05 ± 18.69 | **59 ± 16** |
| | Wet | 4 / 3 | 2.55 ± 3.18 | 4.07 ± 1.40 | 1.74 ± 2.18 | 19.47 ± 21.41 | 0.71 ± 0.41 | 1.34 ± 0.76 | 24.18 ± 2.42 | **51 ± 44** |
| AGL2 (AGL) | Dry | 5 / 4 | 0.67 ± 0.68 | 5.41 ± 3.89 | 1.86 ± 1.25 | 49.85 ± 52.15 | 1.85 ± 2.95 | **1.91 ± 0.77** | 42.45 ± 19.03 | **110 ± 150** |
| | Wet | 4 / 3 | 2.42 ± 4.03 | 3.25 ± 0.72 | 0.96 ± 0.94 | 14.12 ± 12.90 | 0.57 ± 0.23 | 0.99 ± 0.49 | 19.48 ± 2.24 | **230 ± 267** |
| LIV3 (LIV) | Dry | 5 / 4 | 1.10 ± 1.72 | 5.72 ± 3.03 | 2.44 ± 1.85 | 38.74 ± 32.71 | 2.58 ± 4.01 | **3.41 ± 1.68** | 39.32 ± 22.47 | **160 ± 261** |
| | Wet | 4 / 3 | 1.92 ± 1.71 | 3.01 ± 1.80 | 1.85 ± 2.60 | 26.40 ± 16.78 | **3.87 ± 6.14** | **1.57 ± 0.88** | 22.29 ± 5.18 | **55 ± 59** |
| AGL3 (AGL) | Dry | 5 / 4 | 1.73 ± 1.06 | 5.47 ± 4.32 | 3.52 ± 2.59 | 33.35 ± 17.54 | 2.84 ± 3.63 | **1.58 ± 0.38** | 39.55 ± 24.32 | **130 ± 218** |
| | Wet | 4 / 3 | 1.17 ± 1.37 | 2.96 ± 1.70 | 0.89 ± 0.33 | 16.25 ± 11.38 | 0.64 ± 0.09 | 1.11 ± 0.65 | 20.03 ± 2.31 | **26 ± 14** |
| LWL1 (LWL) | Dry | 3 / 2 | 2.31 ± 1.34 | 6.64 ± 5.57 | 9.38 ± 3.40 | 41.39 ± 28.59 | **4.80 ± 4.04** | **3.23 ± 3.29** | 44.09 ± 34.11 | **32 ± 2** |
| | Wet | 4 / 3 | 2.87 ± 2.47 | 4.72 ± 4.36 | 2.45 ± 2.73 | 69.35 ± 24.98 | 0.61 ± 0.37 | 0.84 ± 0.59 | 21.95 ± 4.66 | **104 ± 115** |
| LIV4 (LIV) | Dry | 5 / 4 | 0.68 ± 0.81 | 4.14 ± 1.81 | 2.09 ± 1.16 | 21.74 ± 13.08 | 2.04 ± 1.03 | 1.21 ± 0.42 | 38.19 ± 18.47 | **53 ± 10** |
| | Wet | 4 / 3 | 2.06 ± 2.66 | 3.59 ± 2.86 | 0.90 ± 0.51 | 35.04 ± 29.68 | 0.62 ± 0.40 | 0.82 ± 0.45 | 21.09 ± 1.60 | **96 ± 45** |
| LIV5 (LIV) | Dry | 5 / 4 | 0.85 ± 0.36 | 6.23 ± 3.89 | 3.39 ± 2.82 | 28.53 ± 14.42 | **5.50 ± 5.52** | 1.24 ± 0.26 | 37.51 ± 17.57 | **61 ± 45** |
| | Wet | 4 / 3 | 2.59 ± 3.04 | 2.77 ± 2.08 | 0.72 ± 0.34 | 39.32 ± 59.28 | 1.09 ± 0.72 | 0.84 ± 0.41 | 20.15 ± 3.30 | **70 ± 18** |
| LWL2 (LWL) | Dry | 5 / 4 | 1.06 ± 0.93 | 5.45 ± 5.57 | 2.23 ± 2.59 | 19.48 ± 11.36 | 2.03 ± 2.75 | **1.56 ± 0.34** | 37.53 ± 17.76 | **158 ± 96** |
| | Wet | 4 / 3 | 2.68 ± 2.12 | 3.75 ± 5.34 | 0.84 ± 0.59 | 19.25 ± 17.39 | 0.59 ± 0.29 | 0.75 ± 0.40 | 18.35 ± 2.99 | **227 ± 274** |
| LWL3 (LWL) | Dry | 5 / 4 | 0.57 ± 0.30 | 5.61 ± 6.39 | 2.35 ± 4.67 | 22.02 ± 12.01 | 2.27 ± 4.30 | 1.28 ± 0.30 | 32.97 ± 21.35 | **125 ± 40** |
| | Wet | 4 / 3 | 2.24 ± 3.91 | 1.72 ± 1.39 | 0.30 ± 0.08 | 21.24 ± 33.65 | 0.35 ± 0.18 | 0.98 ± 0.51 | 19.00 ± 0.68 | **169 ± 239** |
| LWL4 (LWL) | Dry | 5 / 4 | 0.77 ± 0.46 | 5.21 ± 5.38 | 1.43 ± 1.44 | 17.65 ± 10.38 | 0.63 ± 0.51 | **1.74 ± 0.26** | 37.04 ± 18.55 | **63 ± 66** |
| | Wet | 4 / 3 | 2.34 ± 2.31 | 2.08 ± 1.37 | 0.75 ± 0.97 | 24.03 ± 32.47 | 1.11 ± 1.05 | 1.16 ± 0.54 | 24.04 ± 10.76 | **73 ± 93** |

NB: Values are mean ± SD of waterpan-visit observations. n, number of sampling visits for nutrients, fluoride and COD / for *E. coli* (totals: 106 / 82). LWL1 was dry in December 2025 and January 2026. A dash indicates no applicable WHO or KEBS guideline. Bold values are above the WHO and/or KEBS guideline; for E. coli, they indicate detectable counts. SRP, soluble reactive phosphorus; TP, total phosphorus; COD, chemical oxygen demand. Land use: AGL, agriculture–livestock; LIV, livestock; LWL, livestock–wildlife.

**Table S4.** Sampling campaigns, dates, season and number of waterpan-visit observations.

| Campaign | Dates | Season | Waterpans sampled | Physicochemical observations | *E. coli* observations | Notes |
|---|---|---|---|---|---|---|
| 1 | 7th–10th November 2024 | Dry | 12 | 12 | — | |
| 2 | 23rd–27th January 2025 | Wet | 12 | 12 | — | |
| 3 | 1st–5th May 2025 | Wet | 12 | 12 | 12 | |
| 4 | 19th–23rd June 2025 | Wet | 12 | 12 | 12 | |
| 5 | 9th–13th October 2025 | Dry | 12 | 12 | 12 | |
| 6 | 19th–24th November 2025 | Dry | 12 | 12 | 12 | |
| 7 | 12th–18th December 2025 | Dry | 11 | 11 | 11 | LWL1 dry |
| 8 | 20th January–1st February 2026 | Dry | 11 | 11 | 11 | LWL1 dry |
| 9 | 26th February–1st March 2026 | Wet | 12 | 12 | 12 | |
| **Total** | | | | **106** | **82** | |

NB: Each waterpan was sampled at three within-pan positions (A, B and C) on every campaign, and the three position values were averaged to give one observation per waterpan per campaign. Season was classified from household-reported rainfall in the 1–2 weeks before sampling. LWL1 was dry in December 2025 and January 2026 and could not be sampled. A dash indicates that *E. coli* was not enumerated on that campaign.

**Table S5.** Effect of waterpan depth on 14 physicochemical water quality parameters, from linear mixed-effects models with waterpan as a random intercept to account for repeated sampling (n = 88 waterpan-occasion observations across 10 waterpans). Depth was not measured at LWL2 and LWL3 because hippopotamuses were present.

| Parameter | β (±SE) | t-value | p-value | n (waterpans) | AIC | $R^2m$ | $R^2c$ |
|---|---|---|---|---|---|---|---|
| Temperature (°C) | -0.317 (±0.501) | -0.63 | 0.539 | 88 (10) | 413.8 | 0.101 | 0.248 |
| pH | 0.120 (±0.120) | 1.00 | 0.322 | 88 (10) | 236.4 | 0.110 | — |
| TDS (mg $L^{-1}$) | -0.042 (±0.041) | -1.02 | 0.328 | 88 (10) | -5.2 | 0.216 | 0.376 |
| EC (µS $cm^{-1}$) | -0.038 (±0.031) | -1.24 | 0.225 | 88 (10) | -86.4 | 0.306 | 0.623 |
| Dissolved $O_2$ (mg $L^{-1}$) | 0.077 (±0.354) | 0.22 | 0.832 | 88 (10) | 410.7 | 0.063 | 0.069 |
| $O_2$ saturation (%) | 1.480 (±5.715) | 0.26 | 0.802 | 88 (10) | 857.9 | 0.058 | 0.081 |
| Secchi depth (cm) | 0.106 (±0.039) | 2.69 | **0.012*** | 88 (10) | -48.0 | 0.289 | 0.623 |
| SRP (µg $L^{-1}$) | 0.015 (±0.093) | 0.16 | 0.873 | 88 (10) | 171.8 | 0.029 | 0.074 |
| TP (mg $L^{-1}$) | -0.010 (±0.049) | -0.21 | 0.837 | 88 (10) | 88.7 | 0.077 | — |
| Nitrate (mg $L^{-1}$) | -0.120 (±0.093) | -1.28 | 0.224 | 88 (10) | 134.7 | 0.263 | 0.396 |
| Ammonium (µg $L^{-1}$) | -0.075 (±0.077) | -0.98 | 0.351 | 88 (10) | 122.5 | 0.054 | 0.150 |
| Nitrite (mg $L^{-1}$) | -0.077 (±0.076) | -1.00 | 0.344 | 88 (10) | 151.8 | 0.174 | 0.191 |
| Fluoride (mg $L^{-1}$) | -0.059 (±0.049) | -1.20 | 0.252 | 88 (10) | 25.3 | 0.237 | 0.383 |
| COD (mg $L^{-1}$) | 0.015 (±0.031) | 0.47 | 0.640 | 88 (10) | 15.7 | 0.189 | — |

NB: β is the standardised coefficient (±SE) for waterpan depth, on the same scale as Table 3: $log_{10}$ for the 10 log-transformed parameters and the measured scale for temperature, pH, dissolved oxygen and oxygen saturation. n is the number of observations, with the number of waterpans in parentheses. $R^2c$ could not be estimated where the waterpan variance was zero. *$p < 0.05$.

**Table S6.** PERMANOVA of water quality composition, by analytical term, based on Euclidean distances from the 14 centred and standardised physicochemical parameters ($log_{10}$-transformed where applicable), with 9,999 permutations matched to the exchangeable unit of each term.

| Term | Analytical level | $R^2$ | F | df | p | Permutation |
|---|---|---|---|---|---|---|
| Waterpan | visits (n = 106) | 0.201 | 2.15 | 11, 94 | **<0.001*** | within campaign |
| Season | visits (n = 106) | 0.096 | 11.08 | 1, 104 | **<0.001*** | strata (waterpan) |
| Season (exact test) | visits (n = 106) | 0.096 | 11.08 | 1 | 0.087 | exact (126 assignments) |
| Land use | waterpan profiles (n = 12) | 0.318 | 2.10 | 2, 9 | 0.051 | free |
| Season × Land use | waterpan shift vectors (n = 12) | 0.207 | 1.17 | 2, 9 | 0.310 | free |

NB: The exact test row is a conservative robustness check on the Season row above it, not an additional independent term. $R^2$ values are comparable only within an analytical level. *$p < 0.05$.

**Table S7.** Homogeneity of multivariate dispersion (betadisper, permutest) for each PERMANOVA term.

| Term | Analytical level | F | p |
|---|---|---|---|
| Waterpan | visits (n = 106) | 1.64 | **0.029*** |
| Season | visits (n = 106) | 0.25 | 0.624 |
| Land use | waterpan profiles (n = 12) | 0.51 | 0.618 |
| Season × Land use | waterpan shift vectors (n = 12) | 0.10 | 0.933 |

NB: *$p < 0.05$, indicating dispersion was not homogeneous for that term.

**Table S8.** Full negative-binomial mixed model results for *E. coli* counts (n = 246 individual samples, nested within 12 waterpans), including random-effects variance components, dispersion, and model-fit statistics (corresponds to Table 4 in the main text, which gives the fixed-effect estimates only).

| **Fixed effect** | **Estimate (SE)** | **z-value** | **p-value** | **IRR (95% CI)** |
|---|---|---|---|---|
| Intercept | 3.833 (0.386) | 9.93 | <0.001*** | 46.19 (21.68–98.41) |
| Season (Dry vs. Wet) | 0.124 (0.463) | 0.27 | 0.788 | 1.13 (0.46–2.81) |
| Land use (Livestock vs. Agriculture-livestock) | 0.069 (0.228) | 0.30 | 0.761 | 1.07 (0.68–1.68) |
| Land use (Livestock-wildlife vs. Agriculture-livestock) | 0.535 (0.243) | 2.20 | 0.028* | 1.71 (1.06–2.75) |
| Waterpan area (standardised) | −0.091 (0.102) | −0.90 | 0.371 | 0.91 (0.75–1.11) |
| **Random effects** | | | | |
| Waterpan (intercept), variance (SD) | 0.018 (0.1353) — 2.4% of random-effects variance | | | |
| Occasion (intercept), variance (SD) | 0.323 (0.5686) — 42.8% of random-effects variance | | | |
| Waterpan:Occasion (intercept), variance (SD) | 0.413 (0.6427) — 54.7% of random-effects variance | | | |
| Dispersion (NB1) | 17.81 | | | |
| **Model fit** | | | | |
| n (individual samples) | 246 | | | |
| n (waterpan-occasion combinations, 12 waterpans, 7 occasions) | 82 | | | |
| df (full model) | 9 | | | |
| log-likelihood (full model) | −1264.713 | | | |
| AIC (full model) | 2547.4 | | | |
| df (null: random effects only) | 5 | | | |
| log-likelihood (null model) | −1266.992 | | | |
| AIC (null model) | 2544.0 | | | |
| ΔAIC (full − null) | +3.43 | | | |
| Likelihood-ratio test (full vs. null) | $\chi^2_4 = 4.57$, p = 0.335 | | | |
| Land use, joint Type II Wald test (2 df) | $\chi^2_2 = 5.80$, p = 0.055 | | | |

NB: Model: *E. coli* ~ Season + Land use + Area + (1 | Waterpan) + (1 | Occasion) + (1 | Waterpan:Occasion) (Equation 3), fitted by maximum likelihood with glmmTMB (Brooks et al., 2017; family = nbinom1, log link); reference categories: Season = Wet, Land use = Agriculture-livestock. Waterpan area is the seasonal mean area of each waterpan, standardised (z-score) prior to fitting. IRR, incidence rate ratio, with 95% Wald confidence intervals. Random-effects variance is partitioned across the waterpan, sampling-occasion and waterpan-occasion (individual visit) levels, because the model was fitted to counts from the three sampling points (A, B and C). ΔAIC is the full model AIC minus the null model AIC. The land-use term was tested with a joint Type II Wald test (car: Anova; Fox & Weisberg, 2019). Significance: * $p < 0.05$, ** $p < 0.01$, *** $p < 0.001$.

**Table S9.** Fluoride exposure and non-carcinogenic hazard by waterpan and season. Values are calculated for each individual sample (RfD = 0.06 mg kg⁻¹ day⁻¹; adults: 2 L day⁻¹, 70 kg; children: 1 L day⁻¹, 15 kg) and summarised as the mean, the maximum (worst case), and the percentage of samples exceeding HQ = 1.

| Waterpan | Season | Land use | n | Fluoride mean (mg L⁻¹) | Child HQ mean | Child HQ max | Child HQ % > 1 | Adult HQ mean | Adult HQ max | Adult HQ % > 1 |
|---|---|---|---|---|---|---|---|---|---|---|
| LIV1 | Dry | LIV | 15 | 2.12 | **2.35** | **3.19** | 100.0 | **1.01** | **1.37** | 60.0 |
| LIV1 | Wet | LIV | 12 | 1.83 | **2.03** | **3.22** | 75.0 | 0.87 | **1.38** | 50.0 |
| AGL1 | Dry | AGL | 15 | 2.65 | **2.95** | **4.09** | 93.3 | **1.26** | **1.75** | 73.3 |
| AGL1 | Wet | AGL | 12 | 2.30 | **2.55** | **4.20** | 91.7 | **1.09** | **1.80** | 50.0 |
| LIV2 | Dry | LIV | 15 | 1.88 | **2.09** | **2.31** | 100.0 | 0.90 | 0.99 | 0.0 |
| LIV2 | Wet | LIV | 12 | 1.34 | **1.49** | **2.62** | 50.0 | 0.64 | **1.12** | 16.7 |
| AGL2 | Dry | AGL | 15 | 1.91 | **2.13** | **3.18** | 86.7 | 0.91 | **1.36** | 53.3 |
| AGL2 | Wet | AGL | 12 | 0.99 | **1.10** | **2.55** | 50.0 | 0.47 | **1.09** | 8.3 |
| LIV3 | Dry | LIV | 15 | 3.41 | **3.79** | **6.05** | 100.0 | **1.62** | **2.59** | 80.0 |
| LIV3 | Wet | LIV | 12 | 1.57 | **1.74** | **2.69** | 75.0 | 0.75 | **1.15** | 50.0 |
| AGL3 | Dry | AGL | 15 | 1.58 | **1.75** | **3.02** | 100.0 | 0.75 | **1.29** | 13.3 |
| AGL3 | Wet | AGL | 12 | 1.11 | **1.24** | **2.02** | 50.0 | 0.53 | 0.87 | 0.0 |
| LWL1 | Dry | LWL | 9 | 3.23 | **3.59** | **8.58** | 88.9 | **1.54** | **3.68** | 55.6 |
| LWL1 | Wet | LWL | 12 | 0.84 | 0.93 | **1.92** | 25.0 | 0.40 | 0.82 | 0.0 |
| LIV4 | Dry | LIV | 15 | 1.21 | **1.34** | **1.85** | 73.3 | 0.57 | 0.79 | 0.0 |
| LIV4 | Wet | LIV | 12 | 0.82 | 0.91 | **1.50** | 41.7 | 0.39 | 0.64 | 0.0 |
| LIV5 | Dry | LIV | 15 | 1.24 | **1.38** | **1.65** | 86.7 | 0.59 | 0.71 | 0.0 |
| LIV5 | Wet | LIV | 12 | 0.84 | 0.93 | **1.42** | 50.0 | 0.40 | 0.61 | 0.0 |
| LWL2 | Dry | LWL | 15 | 1.56 | **1.73** | **2.24** | 93.3 | 0.74 | 0.96 | 0.0 |
| LWL2 | Wet | LWL | 12 | 0.75 | 0.83 | **1.50** | 25.0 | 0.36 | 0.64 | 0.0 |
| LWL3 | Dry | LWL | 15 | 1.28 | **1.42** | **1.83** | 80.0 | 0.61 | 0.78 | 0.0 |
| LWL3 | Wet | LWL | 12 | 0.98 | **1.09** | **1.66** | 50.0 | 0.47 | 0.71 | 0.0 |
| LWL4 | Dry | LWL | 15 | 1.74 | **1.93** | **2.38** | 100.0 | 0.83 | **1.02** | 6.7 |
| LWL4 | Wet | LWL | 12 | 1.16 | **1.29** | **2.18** | 50.0 | 0.55 | 0.94 | 0.0 |

NB: n is lower in the dry season at LWL1 because the pan held no standing water to sample in December 2025 and January 2026; all other differences in n reflect the five dry and four wet sampling campaigns. Bold indicates HQ ≥ 1. AGL, agriculture-livestock; LIV, livestock; LWL, livestock-wildlife. Values pooled across seasons are given in Table S10.

**Table S10**. Fluoride exposure and non-carcinogenic hazard by waterpan, pooled across both seasons. Values are calculated for each individual sample and summarised per waterpan (n = 27 samples per waterpan; 21 for LWL1). CDI = C × IR / BW; HQ = CDI / RfD, with RfD = 0.06 mg kg⁻¹ day⁻¹. Adults: 2 L day⁻¹, 70 kg. Children: 1 L day⁻¹, 15 kg. HQ max is the highest sample-level hazard quotient at that waterpan, i.e. the worst case.

| Waterpan | Land use | n | Fluoride mean ± SD (mg L⁻¹) | Fluoride max (mg L⁻¹) | Adult HQ mean | Adult HQ max | Child HQ mean | Child HQ max | Adult HQ % > 1 | Child HQ % > 1 |
|---|---|---|---|---|---|---|---|---|---|---|
| LIV1 | LIV | 27 | 1.99 ± 0.78 | 2.90 | 0.95 | **1.38** | **2.21** | **3.22** | 55.6 | 88.9 |
| AGL1 | AGL | 27 | 2.49 ± 1.07 | 3.78 | **1.19** | **1.80** | **2.77** | **4.20** | 63.0 | 92.6 |
| LIV2 | LIV | 27 | 1.64 ± 0.55 | 2.36 | 0.78 | **1.12** | **1.82** | **2.62** | 7.4 | 77.8 |
| AGL2 | AGL | 27 | 1.50 ± 0.81 | 2.86 | 0.72 | **1.36** | **1.67** | **3.18** | 33.3 | 70.4 |
| LIV3 | LIV | 27 | 2.59 ± 1.56 | 5.44 | **1.23** | **2.59** | **2.88** | **6.05** | 66.7 | 88.9 |
| AGL3 | AGL | 27 | 1.37 ± 0.56 | 2.72 | 0.65 | **1.29** | **1.52** | **3.02** | 7.4 | 77.8 |
| LWL1 | LWL | 21 | 1.86 ± 2.25 | 7.73 | 0.89 | **3.68** | **2.07** | **8.58** | 23.8 | 52.4 |
| LIV4 | LIV | 27 | 1.04 ± 0.45 | 1.67 | 0.49 | 0.79 | **1.15** | **1.85** | 0.0 | 59.3 |
| LIV5 | LIV | 27 | 1.06 ± 0.37 | 1.49 | 0.51 | 0.71 | **1.18** | **1.65** | 0.0 | 70.4 |
| LWL2 | LWL | 27 | 1.20 ± 0.54 | 2.02 | 0.57 | 0.96 | **1.33** | **2.24** | 0.0 | 63.0 |
| LWL3 | LWL | 27 | 1.14 ± 0.39 | 1.65 | 0.55 | 0.78 | **1.27** | **1.83** | 0.0 | 66.7 |
| LWL4 | LWL | 27 | 1.48 ± 0.47 | 2.14 | 0.71 | **1.02** | **1.65** | **2.38** | 3.7 | 77.8 |

NB: Bold indicates HQ ≥ 1. Seasonally disaggregated values are given in Table S9. AGL, agriculture-livestock; LIV, livestock; LWL, livestock-wildlife.

**Table S11.** Full linear mixed-effects model results for the 14 physicochemical water quality parameters, including random-effects variance components, AIC and $R^2$ (corresponds to Table 3 in the main text, which gives the fixed-effect estimates only).

| Fixed effects | Temperature (°C) | pH | TDS (mg L⁻¹) | EC (µS cm⁻¹) | Dissolved $O_2$ (mg L⁻¹) | $O_2$ saturation (%) | Secchi depth (cm) |
|---|---|---|---|---|---|---|---|
| Intercept | 20.79 (±0.830) 25.05*** | 8.58 (±0.192) 44.72*** | 2.05 (±0.060) 33.90*** | 2.19 (±0.055) 40.01*** | 5.05 (±0.515) 9.81*** | 74.31 (±8.485) 8.76*** | 0.72 (±0.109) 6.61*** |
| Season (Dry) | 0.10 (±0.456) 0.22 | 0.65 (±0.170) 3.81*** | 0.17 (±0.035) 4.90*** | 0.15 (±0.021) 7.03*** | -0.71 (±0.456) -1.55 | -10.25 (±6.996) -1.46 | 0.01 (±0.034) 0.29 |
| Land use (LIV) | 1.72 (±1.020) 1.68 | -0.05 (±0.216) -0.24 | -0.06 (±0.074) -0.80 | -0.06 (±0.069) -0.84 | -0.43 (±0.578) -0.74 | -6.46 (±9.736) -0.66 | 0.06 (±0.138) 0.40 |
| Land use (LWL) | 1.88 (±1.076) 1.74 | 0.30 (±0.230) 1.31 | 0.12 (±0.078) 1.58 | 0.13 (±0.072) 1.76 | 0.69 (±0.617) 1.12 | 11.17 (±10.365) 1.08 | 0.24 (±0.145) 1.62 |
| Area | -0.14 (±0.440) -0.33 | -0.08 (±0.095) -0.85 | -0.02 (±0.032) -0.54 | -0.02 (±0.029) -0.58 | 0.07 (±0.255) 0.29 | 0.99 (±4.280) 0.23 | 0.06 (±0.057) 0.99 |
| Waterpan SD | 1.134 | 0 | 0.08 | 0.086 | 0 | 5.386 | 0.178 |
| Residual SD | 2.298 | 0.868 | 0.177 | 0.104 | 2.326 | 35.675 | 0.165 |
| AIC | 496.2 | 290.3 | -22.7 | -123.1 | 489.5 | 1042.4 | -25.3 |
| $R^2$m | 0.088 | 0.144 | 0.25 | 0.375 | 0.070 | 0.067 | 0.215 |
| $R^2$c | 0.266 | — | 0.376 | 0.628 | — | 0.088 | 0.636 |
| **Fixed effects** | **SRP (µg L⁻¹)** | **TP (mg L⁻¹)** | **Soluble nitrate (mg L⁻¹)** | **Soluble ammonium (µg L⁻¹)** | **Soluble nitrite (mg L⁻¹)** | **Fluoride (mg L⁻¹)** | **COD (mg L⁻¹)** |
| Intercept | 0.03 (±0.132) 0.20 | 0.46 (±0.080) 5.68*** | -0.05 (±0.169) -0.31 | 1.27 (±0.119) 10.69*** | -0.19 (±0.111) -1.74 | 0.05 (±0.078) 0.69 | 1.29 (±0.045) 28.78*** |
| Season (Dry) | -0.12 (±0.104) -1.16 | 0.18 (±0.071) 2.57* | 0.47 (±0.088) 5.32*** | 0.16 (±0.080) 1.95 | 0.36 (±0.098) 3.63*** | 0.25 (±0.041) 5.93*** | 0.20 (±0.040) 4.94*** |
| Land use (LIV) | -0.05 (±0.154) -0.35 | -0.06 (±0.090) -0.69 | -0.05 (±0.209) -0.24 | -0.06 (±0.143) -0.42 | 0.14 (±0.124) 1.12 | -0.03 (±0.096) -0.29 | 0.02 (±0.051) 0.44 |
| Land use (LWL) | 0.04 (±0.163) 0.22 | -0.14 (±0.096) -1.40 | -0.24 (±0.220) -1.07 | -0.09 (±0.151) -0.61 | -0.22 (±0.133) -1.65 | -0.14 (±0.101) -1.38 | 0.02 (±0.054) 0.35 |
| Area | 0.02 (±0.067) 0.29 | -0.03 (±0.040) -0.68 | -0.03 (±0.090) -0.28 | -0.02 (±0.062) -0.33 | 0.03 (±0.055) 0.52 | 0.04 (±0.041) 1.00 | -0.02 (±0.022) -1.00 |
| Waterpan SD | 0.107 | 0 | 0.238 | 0.135 | 0 | 0.108 | 0 |
| Residual SD | 0.529 | 0.363 | 0.444 | 0.408 | 0.501 | 0.208 | 0.203 |
| AIC | 192.6 | 114.2 | 165.0 | 143.0 | 179.2 | 12.0 | -2.8 |
| $R^2$m | 0.022 | 0.091 | 0.214 | 0.043 | 0.174 | 0.244 | 0.205 |
| $R^2$c | 0.061 | — | 0.390 | 0.138 | — | 0.406 | — |

NB: Fixed effects are given as coefficient β (±SE) and t-value. Coefficients are on the $\log_{10}$ scale for the ten log-transformed parameters and on the measured scale for temperature, pH, dissolved oxygen and oxygen saturation, which were analysed untransformed. AIC is therefore comparable only within a parameter, never across parameters on different scales. Reference levels: Season = Wet, Land use

= Agriculture–livestock, so a positive season coefficient indicates a higher value in the dry season. $R^2m$, marginal $R^2$ (fixed effects only); $R^2c$, conditional $R^2$ (fixed + random effects); unavailable where the waterpan random effect was estimated at zero. Significance: *$p<0.05$, **$p<0.01$, ***$p<0.001$.

**Table S12**. Water quality parameters across all waterpans and sampling occasions, and by season and land use category, compared with WHO (2022) and KEBS (2014) drinking water guideline values. Parameters without numerical guideline are reported descriptively.

| Parameter | Guideline (WHO / KEBS) | Overall | | | | Dry season | | | Wet season | | |
|---|---|---|---|---|---|---|---|---|---|---|---|
| | | n | Mean ± SD | Median | Range (min–max) | AGL | LIV | LWL | AGL | LIV | LWL |
| *n (physicochemical / E. coli)* | | | | | | *15 / 12* | *25 / 20* | *18 / 14* | *12 / 9* | *20 / 15* | *16 / 12* |
| Temperature (°C) | – | 106 | 22.19 ± 2.57 | 21.95 | 14.60–28.67 | 20.51 ± 3.46 | 22.87 ± 1.99 | 22.82 ± 2.46 | 21.26 ± 2.50 | 22.32 ± 2.53 | 22.50 ± 2.04 |
| pH | 6.5–8.5 / 5.5–9.5 | 106 | **9.01 ± 0.92** | 9.25 | 7.30–12.29 | **9.11 ± 0.69** | **9.14 ± 0.69** | **9.70 ± 1.09** | **8.73 ± 0.88** | **8.66 ± 0.95** | **8.59 ± 0.81** |
| TDS (mg $L^{-1}$) | 1000 / 1500 | 106 | 161.12 ± 81.32 | 147.00 | 30.00–562.67 | 172.40 ± 64.26 | 153.50 ± 56.85 | 247.69 ± 110.96 | 125.04 ± 51.35 | 111.60 ± 46.17 | 154.04 ± 71.39 |
| EC (µS $cm^{-1}$) | – / 2500 | 106 | 206.31 ± 76.94 | 192.17 | 69.00–563.33 | 215.94 ± 49.91 | 199.16 ± 66.27 | 307.20 ± 76.04 | 166.16 ± 41.04 | 145.77 ± 37.08 | 200.75 ± 62.90 |
| Dissolved $O_2$ (mg $L^{-1}$) | – | 106 | 4.70 ± 2.37 | 4.45 | 0.93–11.90 | 4.41 ± 2.86 | 3.68 ± 1.90 | 5.29 ± 2.04 | 4.97 ± 2.68 | 4.84 ± 2.47 | 5.55 ± 2.26 |
| $O_2$ saturation (%) | – | 106 | 69.57 ± 36.58 | 66.12 | 17.23–187.67 | 64.53 ± 45.43 | 54.62 ± 28.40 | 78.88 ± 32.80 | 73.78 ± 41.61 | 70.63 ± 37.37 | 82.71 ± 34.47 |
| Secchi depth (cm) | – | 106 | 8.30 ± 6.50 | 6.00 | 2.50–31.67 | 4.95 ± 1.56 | 5.85 ± 2.45 | 15.74 ± 9.81 | 6.73 ± 2.66 | 6.48 ± 2.22 | 10.38 ± 8.05 |
| SRP (µg $L^{-1}$) | – / 2200 | 106 | 1.69 ± 2.00 | 0.98 | 0.03–8.49 | 1.26 ± 0.87 | 1.06 ± 0.96 | 1.05 ± 0.92 | 2.20 ± 2.96 | 2.37 ± 2.69 | 2.53 ± 2.51 |
| Total phosphorus (mg $L^{-1}$) | – | 106 | 4.29 ± 3.46 | 2.94 | 0.42–15.26 | 5.25 ± 3.54 | 4.94 ± 2.96 | 5.63 ± 5.25 | 3.53 ± 1.78 | 3.01 ± 1.91 | 3.07 ± 3.44 |
| Soluble nitrate (mg $L^{-1}$) | 50 / 45 | 106 | 2.44 ± 2.82 | 1.31 | 0.07–13.25 | 4.04 ± 3.33 | 3.21 ± 2.47 | 3.23 ± 4.09 | 1.40 ± 1.83 | 1.29 ± 1.49 | 1.09 ± 1.57 |
| Soluble ammonium (µg $L^{-1}$) | – / 500 | 106 | 29.95 ± 26.23 | 22.24 | 0.86–136.71 | 39.20 ± 32.37 | 32.40 ± 22.80 | 23.33 ± 16.05 | 24.58 ± 17.98 | 26.29 ± 30.68 | 33.47 ± 32.92 |
| Soluble nitrite (mg $L^{-1}$) | 3 / 3 | 106 | 2.19 ± 3.15 | 0.76 | 0.07–13.42 | **3.34 ± 3.62** | **3.82 ± 3.94** | 2.17 ± 3.17 | 0.66 ± 0.20 | 1.44 ± 2.78 | 0.67 ± 0.60 |
| Fluoride (mg $L^{-1}$) | 1.5 / 1.5 | 106 | **1.61 ± 1.03** | 1.39 | 0.29–6.99 | **2.05 ± 0.71** | **1.97 ± 1.11** | **1.81 ± 1.34** | 1.47 ± 1.07 | 1.28 ± 0.80 | 0.93 ± 0.49 |
| COD (mg $L^{-1}$) | – | 106 | 29.44 ± 16.80 | 22.53 | 4.80–71.87 | 38.09 ± 21.57 | 37.65 ± 17.89 | 37.27 ± 20.32 | 19.17 ± 2.06 | 21.36 ± 3.43 | 20.84 ± 5.91 |
| *E. coli* (CFU 100 $mL^{-1}$) | Not detectable in 100 mL | 82 | **95 ± 122** | 51 | 9–550 | **96 ± 144** | **85 ± 115** | **103 ± 76** | **94 ± 169** | **63 ± 40** | **144 ± 179** |

NB: Values are mean ± SD of waterpan-visit observations. n, number of waterpan-visit observations; in the season × land use columns, given as physicochemical */ E. coli.* A dash indicates no applicable WHO or KEBS guideline. Bold values are above the WHO and/or KEBS guideline; for pH, above the WHO upper limit of 8.5; for *E. coli*, they indicate detectable counts. TDS, total dissolved solids; EC, electrical conductivity; SRP, soluble reactive phosphorus; COD, chemical oxygen demand. Land use: AGL, agriculture–livestock; LIV, livestock; LWL, livestock–wildlife.

**Table S13.** *E. coli* concentrations by waterpan and season, expressed as WHO (1997) drinking water risk categories assigned to individual samples: in conformity (0 CFU 100 mL$^{-1}$), low (1–10), intermediate (11–100), high (101–1000), very high (> 1000). No sample fell in the very-high category, which is therefore omitted. Non-compliance is the percentage of samples with any detectable *E. coli*. Counts in the high category are shown in bold.

| | | | | *E. coli* (CFU 100 mL$^{-1}$) | | | | | | | | |
|---|---|---|---|---|---|---|---|---|---|---|---|---|
| **Waterpan** | **Land use** | **Season** | ***n*** | **Mean** | **Median** | **Range** | **In conformity** | **Low** | **Intermediate** | **High** | **% high** | **% non-compliant** |
| LIV1 | LIV | Dry | 12 | 91.1 | 58.5 | 30–246 | 0 | 0 | 7 | **5** | 41.7 | 100.0 |
| LIV1 | LIV | Wet | 9 | 41.7 | 30.0 | 1–125 | 0 | 2 | 6 | **1** | 11.1 | 100.0 |
| AGL1 | AGL | Dry | 12 | 48.8 | 46.0 | 9–144 | 0 | 1 | 10 | **1** | 8.3 | 100.0 |
| AGL1 | AGL | Wet | 9 | 27.0 | 15.0 | 4–92 | 0 | 3 | 6 | **0** | 0.0 | 100.0 |
| LIV2 | LIV | Dry | 12 | 58.8 | 52.0 | 32–95 | 0 | 0 | 12 | **0** | 0.0 | 100.0 |
| LIV2 | LIV | Wet | 9 | 50.6 | 46.0 | 5–139 | 0 | 2 | 6 | **1** | 11.1 | 100.0 |
| AGL2 | AGL | Dry | 12 | 109.9 | 34.0 | 17–388 | 0 | 0 | 9 | **3** | 25.0 | 100.0 |
| AGL2 | AGL | Wet | 9 | 230.4 | 74.0 | 20–599 | 0 | 0 | 5 | **4** | 44.4 | 100.0 |
| LIV3 | LIV | Dry | 12 | 159.9 | 30.5 | 8–656 | 0 | 2 | 7 | **3** | 25.0 | 100.0 |
| LIV3 | LIV | Wet | 9 | 54.6 | 28.0 | 9–130 | 0 | 1 | 5 | **3** | 33.3 | 100.0 |
| AGL3 | AGL | Dry | 12 | 129.5 | 21.5 | 3–626 | 0 | 3 | 6 | **3** | 25.0 | 100.0 |
| AGL3 | AGL | Wet | 9 | 25.7 | 21.0 | 0–63 | 1 | 1 | 7 | **0** | 0.0 | 88.9 |
| LWL1 | LWL | Dry | 6 | 31.8 | 29.0 | 15–58 | 0 | 0 | 6 | **0** | 0.0 | 100.0 |
| LWL1 | LWL | Wet | 9 | 103.9 | 43.0 | 11–306 | 0 | 0 | 6 | **3** | 33.3 | 100.0 |
| LIV4 | LIV | Dry | 12 | 53.0 | 46.5 | 5–170 | 0 | 1 | 10 | **1** | 8.3 | 100.0 |
| LIV4 | LIV | Wet | 9 | 96.1 | 91.0 | 15–195 | 0 | 0 | 5 | **4** | 44.4 | 100.0 |
| LIV5 | LIV | Dry | 12 | 60.7 | 42.5 | 8–184 | 0 | 1 | 9 | **2** | 16.7 | 100.0 |
| LIV5 | LIV | Wet | 9 | 70.3 | 68.0 | 40–102 | 0 | 0 | 7 | **2** | 22.2 | 100.0 |
| LWL2 | LWL | Dry | 12 | 157.6 | 134.5 | 29–329 | 0 | 0 | 4 | **8** | 66.7 | 100.0 |
| LWL2 | LWL | Wet | 9 | 227.3 | 107.0 | 7–609 | 0 | 1 | 3 | **5** | 55.6 | 100.0 |
| LWL3 | LWL | Dry | 12 | 125.2 | 106.5 | 15–263 | 0 | 0 | 6 | **6** | 50.0 | 100.0 |
| LWL3 | LWL | Wet | 9 | 169.4 | 40.0 | 5–470 | 0 | 1 | 5 | **3** | 33.3 | 100.0 |
| LWL4 | LWL | Dry | 12 | 63.2 | 40.5 | 7–164 | 0 | 2 | 7 | **3** | 25.0 | 100.0 |
| LWL4 | LWL | Wet | 9 | 73.4 | 33.0 | 6–211 | 0 | 2 | 4 | **3** | 33.3 | 100.0 |

NB: n, number of individual samples (sampling points A, B and C across visits); n differs between waterpans and seasons because *E. coli* was not measured on the first two sampling occasions and LWL1 was dry in December 2025 and January 2026. AGL, agriculture–livestock; LIV, livestock; LWL, livestock–wildlife.

**Fig. S1** Time series of pH, fluoride, nitrite and *E. coli* in each waterpan across the nine sampling occasions

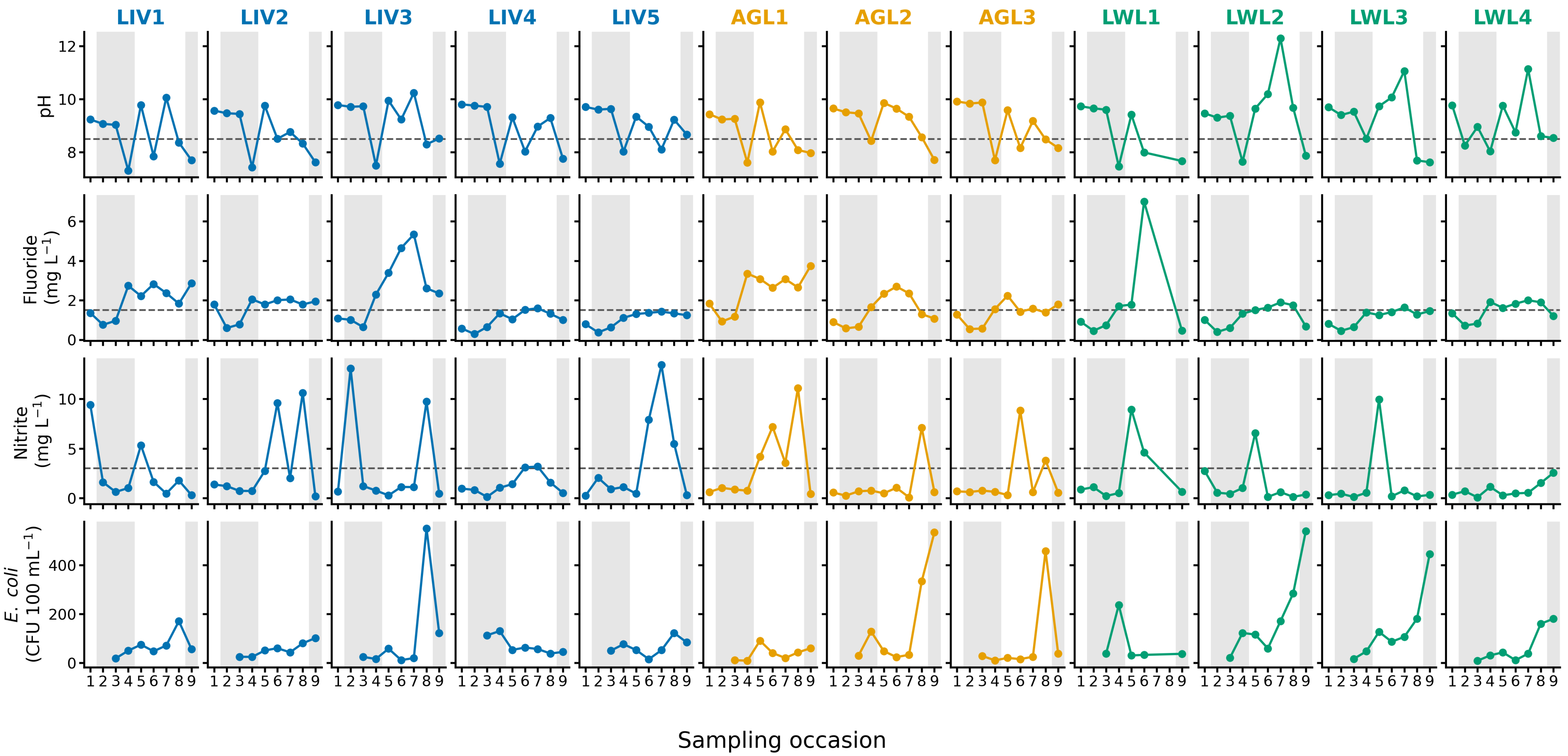


NB: Values are visit means of the three sampling points (A, B and C). Sampling occasions: 1, November 2024; 2, January 2025; 3, May 2025; 4, June 2025; 5, October 2025; 6, November 2025; 7, December 2025; 8, January 2026; 9, February 2026. Grey shading marks wet-season occasions, and dashed lines show WHO/KEBS guideline values. *E. coli* was measured from May 2025 onwards, and LWL1 was dry and not sampled in December 2025 and January 2026. Land use: AGL, agriculture–livestock; LIV, livestock; LWL, livestock–wildlife.
